# Prospective Comparison of [^68^Ga]Ga-FAPI-04 PET, [^18^F]FDG PET, and Contrast-Enhanced MRI for Predicting Pathologic Response after Neoadjuvant Chemotherapy in Breast Cancer

**DOI:** 10.64898/2026.05.05.26352015

**Authors:** Yuming Luo, Xiao Zhang, Runze Li, Yalan Zeng, Yu Zhao, Lei Li, Bei Qian, Yunxiao Xiao, Mengting Li, Yaqi Zhao, Silin Xu, Qin Yang, Huanwei Zhang, Hengyu Chen, Chong Lu, Xiaoli Lan, Chunping Liu

## Abstract

Assessment of pathologic complete response (pCR) following neoadjuvant chemotherapy (NAC) remains an unmet clinical need in breast cancer. Fibroblast activation protein inhibitor (FAPI) PET targets the tumor microenvironment and may therefore enhance response evaluation after NAC. This study aimed to compare the performance of [^68^Ga]Ga-FAPI-04 PET, [^18^F]FDG PET, and contrast-enhanced MRI for predicting pathologic response after NAC in breast cancer, with separate analyses for primary breast lesions and axillary lymph nodes.

**Methods:** In this prospective single-center diagnostic accuracy study, women with biopsy-confirmed stage II-III breast cancer underwent baseline and post-therapy [^68^Ga]Ga-FAPI-04 PET/MRI, [^18^F]FDG PET/CT, and contrast-enhanced MRI before surgery. Quantitative PET parameters were evaluated for primary tumors and axillary lymph nodes. pCR was defined as ypT0/isN0. Significant variables identified in univariable analyses were further explored using least absolute shrinkage and selection operator (LASSO) analysis, and receiver-operating-characteristic (ROC) analysis was performed to assess diagnostic performance. Fibroblast activation protein expression was also assessed by immunohistochemistry in paired pre- and post-therapy tumor specimens from a subset of patients.

**Results:** Twenty-four patients completed the study protocol, yielding 25 primary lesions and 44 metastatic lymph nodes across 27 axillary compartments. Overall patient-level pCR was achieved in 13 of 24 patients (54.17%). The lesion-level pCR rate was 60.00% (15/25) for primary breast lesions, and the node-level pCR rate was 72.73% (32/44) for axillary lymph nodes. For primary tumor response, post-therapy [^68^Ga]Ga-FAPI-04 SUVmax showed the highest diagnostic performance (AUC, 0.84; sensitivity, 80.00%; specificity, 80.00%; accuracy, 80.00%), whereas the optimal [^18^F]FDG parameter was Δ TBR% (AUC, 0.747). For nodal response, post-therapy [^68^Ga]Ga-FAPI-04 SULmean showed the highest diagnostic performance (AUC, 0.89; sensitivity, 91.67%; specificity, 81.25%; accuracy, 84.09%) and was significantly different from the best [^18^F]FDG parameter (Δ SULmax%, AUC, 0.669) on DeLong testing (P < 0.05). MRI achieved AUCs of 0.733 for primary lesions and 0.770 for lymph nodes. Stromal FAP expression positively correlated with [^68^Ga]Ga-FAPI-04 SUVmax and was markedly reduced in lesions achieving pCR.

**Conclusion:** Post-therapy [^68^Ga]Ga-FAPI-04 PET may serve as a promising adjunctive imaging biomarker for predicting pathologic response after NAC in breast cancer, particularly for axillary nodal assessment. These findings suggest that FAPI PET may provide clinically relevant information for preoperative evaluation of residual disease burden, potentially contributing to more individualized surgical planning and treatment decision-making.

## INTRODUCTION

Neoadjuvant chemotherapy (NAC) has become a standard treatment approach for patients with biologically aggressive or locally advanced breast cancer, as it enables tumor downstaging, increases the likelihood of breast-conserving surgery, and allows in vivo assessment of treatment sensitivity (1). Notably, achieving pathologic complete response (pCR) after NAC is associated with improved long-term outcomes, especially in HER2-positive and triple-negative subtypes, underscoring the clinical importance of accurate response assessment for both prognostication and subsequent therapeutic decision-making (2). Recent NeoSTEEP recommendations have reinforced standardized definitions of pCR in neoadjuvant breast cancer trials, emphasizing consistent reporting of residual disease in both the breast and regional lymph nodes (3).

Breast MRI is currently the most established imaging modality for monitoring treatment response after NAC due to its high soft-tissue contrast and sensitivity for detecting residual enhancing tumor (4). However, its performance is suboptimal and may vary according to molecular subtype, treatment regimen, and imaging characteristics such as background parenchymal enhancement or residual nonmass enhancement. Moreover, false-positive findings —attributable to treatment-related fibrosis, inflammation, or isolated residual ductal carcinoma in situ—can compromise specificity (5).

[^18^F]FDG PET provides complementary biological information by measuring glucose metabolism and has demonstrated utility in assessing treatment response in breast cancer, including the prediction of pCR (6). FDG-based imaging also contributes to response-adapted therapeutic strategies, particularly in HER2-positive disease (7).

Fibroblast activation protein (FAP) is highly expressed in cancer-associated fibroblasts within the tumor stroma and plays a critical role in extracellular matrix remodeling, tumor invasion, immune modulation, and treatment resistance (8). In breast cancer, accumulating evidence indicates that FAP-positive stromal cells are associated with biologically aggressive tumor phenotypes and poor clinical outcomes (9). These biological characteristics provide a strong rationale for FAP-targeted molecular imaging. Compared with [^18^F]FDG, radiolabeled fibroblast activation protein inhibitors (FAPIs) typically demonstrate superior tumor-to-background contrast and may more accurately reflect stromal changes following therapy (10).

Preliminary studies of [^68^Ga]Ga-FAPI PET/MRI in breast cancer have demonstrated promising performance in assessing response to neoadjuvant chemotherapy (NAC), with excellent lesion detectability and strong correlation with stromal FAP expression (11). More recent investigations in breast cancer and broader diagnostic evaluations further suggest that FAP-targeted imaging may surpass or complement conventional modalities, particularly in scenarios requiring high lesion-to-background contrast (12,13). However, prospective head-to-head comparisons of [^68^Ga]Ga-FAPI-04, [^18^F]FDG, and contrast-enhanced MRI for predicting pathologic complete response (pCR) in both primary tumors and axillary lymph nodes remain limited.

Therefore, the objective of this prospective study was to evaluate and compare the diagnostic performance of [^68^Ga]Ga-FAPI-04 PET/MRI, [^18^F]FDG PET/CT, and contrast-enhanced MRI in predicting pathologic response to NAC in patients with breast cancer, with separate assessments for primary breast lesions and axillary lymph nodes, and to investigate the histopathologic correlates of FAPI uptake using FAP immunohistochemistry.

## MATERIALS AND METHODS

### Patients

This prospective, single-center diagnostic accuracy study was approved by the Ethics Committee of Union Hospital, Tongji Medical College, Huazhong University of Science and Technology. This study was registered at ClinicalTrials.gov (NCT07553741). Between April 2025 and December 2025, 27 consecutive patients with biopsy-confirmed breast cancer scheduled to receive NAC were enrolled. Written informed consent was obtained from all participants before study inclusion.

Eligible patients were aged 18–70 years, had pathologically confirmed primary breast cancer with TNM stage T2–4N0M0 or T0–4N1–3M0, had not received prior antitumor therapy, had no severe hematologic, cardiac, pulmonary, hepatic, or renal dysfunction, had no immunodeficiency, and had an Eastern Cooperative Oncology Group (ECOG) performance status of 0 or 1. Women of childbearing potential were required to use effective contraception. Exclusion criteria included refusal to undergo [^68^Ga]Ga-FAPI-04 PET/MRI, contraindications to chemotherapy or surgery, presence of distant metastasis, history of prior malignancy or antitumor treatment, hypersensitivity to monoclonal antibodies, autoimmune disorders, active infection, psychiatric illness or cognitive impairment, and pregnancy or lactation.

### Sample Size Estimation

Sample size estimation for this prospective exploratory diagnostic accuracy study was based on receiver-operating characteristic (ROC) analysis for the primary endpoint-discrimination of pCR after NAC. Assuming an expected area under the ROC curve (AUC) of 0.80–0.85, a two-sided α level of 0.05, and 80% statistical power, the minimum required number of evaluable patients was estimated to be 20–24. To account for potential patient dropout or incomplete imaging or pathological data, the target enrollment was set at 27 patients.

### Procedures, NAC Protocols, and Surgical Methods

Following enrollment, all patients underwent baseline [^68^Ga]Ga-FAPI-04 PET/MRI, [^18^F]FDG PET/CT, and contrast-enhanced MRI, with initiation of NAC within 1 week after imaging. Core needle biopsy specimens were analyzed by immunohistochemistry for hormone receptor and HER2 status to determine molecular subtype. Patients were classified as luminal/HER2-negative, HER2-positive, or triple-negative. Patients received molecular subtype-guided NAC regimens according to institutional practice and the China Anti-Cancer Association guidelines. HER2-positive tumors were treated with docetaxel, carboplatin, trastuzumab, and pertuzumab (TCbHP) for 6 cycles. Triple-negative tumors were treated with epirubicin plus cyclophosphamide followed by paclitaxel (EC-T) combined with anti–programmed cell death protein 1 (PD-1) immunotherapy for 8 cycles. Luminal/HER2-negative tumors were treated with epirubicin plus cyclophosphamide followed by paclitaxel (EC-T) for 8 cycles. Drug dosing was administered in accordance with the China Anti-Cancer Association guidelines (14).

At 2-3 weeks after completion of the final NAC cycle, all 3 imaging examinations were repeated, followed by surgery within 1 week (Fig. 1). Sixteen patients underwent modified radical mastectomy, and 8 underwent breast-conserving surgery. Axillary lymph node dissection was performed in 19 patients, whereas 5 patients without baseline axillary nodal metastasis underwent sentinel lymph node biopsy alone. Surgical specimens were serially sectioned and evaluated histopathologically and by immunohistochemistry. Biopsy and surgical specimens also underwent semiquantitative fibroblast activation protein (FAP) immunohistochemical staining. Stromal FAP expression was scored according to the proportion of FAP-positive stromal area as follows: 0, less than 1%; 1, 1%-10%; 2, 11%-50%; and 3, greater than 50% (15,16). Axillary lymph nodes were classified into levels I, II, and III according to their anatomic relationship to the pectoralis minor muscle (17). Pathologic complete response (pCR) was defined as the absence of residual invasive carcinoma in both the breast and axillary lymph nodes, irrespective of the presence of residual in situ disease in the breast (ypT0/isN0M0) (3).

**FIGURE 1.**
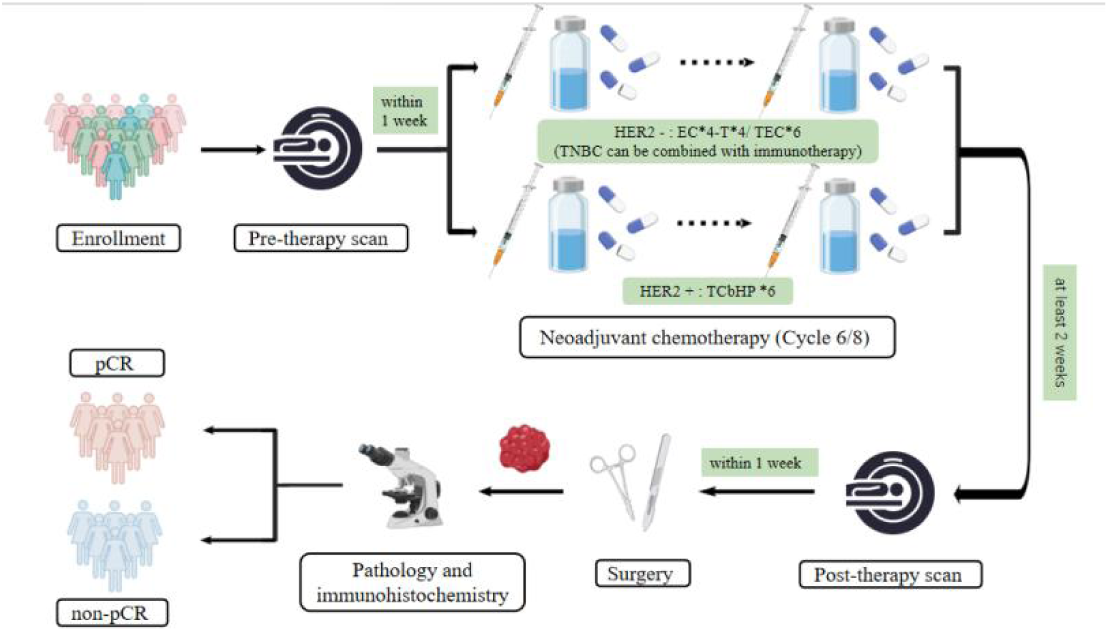
Procedures of clinical trial.

### [^68^Ga]Ga-FAPI-04 PET/MRI, [^18^F]FDG PET/CT, and Contrast-Enhanced MRI Acquisition

Patients underwent breast and axillary PET/MRI approximately 30 minutes after intravenous injection of 1.8–2.2 MBq/kg of [^68^Ga]Ga-FAPI-04. Imaging was performed in the prone position with both breasts suspended naturally within a dedicated breast coil, using an acquisition time of 8 minutes per bed position. [^18^F]FDG PET/CT was conducted 60 minutes after intravenous administration of 3.7–5.5 MBq/kg of [^18^F]FDG using a PET/CT scanner (Discovery VCT; GE Healthcare). Contrast-enhanced MRI was performed on a 3.0-T system (Philips Ingenia) and included T1-weighted, T2-weighted, and diffusion-weighted imaging sequences. The interval between any two imaging examinations was at least 24 hours.

### Image Interpretation

PET images were independently reviewed by 2 nuclear medicine physicians using PET VCAR software (GE Healthcare). Volumes of interest were automatically delineated and manually adjusted when necessary, and discrepancies were resolved by a 3rd experienced physician. Quantitative PET parameters included SUVmax, SUVmean, SULmax, SULmean, SULpeak, MTV, FTV, TLG, TLF, and TBR. TLG and TLF were calculated as SUVmean x MTV and SUVmean x FTV, respectively. TBR was defined as the ratio of lesion SUVmean to the SUVmean of contralateral normal breast tissue. Percentage changes between baseline and post-therapy scans were calculated for SUVmax, SULmax, SULpeak, MTV/FTV, TLG/TLF, and TBR. Lesion-level analysis was performed for breast primaries. Because 1 patient had occult breast cancer without an identifiable breast lesion and 2 patients had 2 spatially distinct breast lesions, 25 primary lesions were included in lesion-level analyses.

All imaging assessments were performed in a blinded manner. The nuclear medicine physicians who reviewed the [^68^Ga]Ga-FAPI-04 PET and [^18^F]FDG PET images were blinded to histopathologic results, clinical outcome data, and the findings of the other imaging modalities. Likewise, contrast-enhanced MRI was interpreted independently without access to PET findings or pathologic results. Pathologic evaluation was also conducted without knowledge of the imaging findings.

For contrast-enhanced MRI, treatment response was assessed on the basis of changes in tumor size, apparent diffusion coefficient (ADC), and residual enhancement in the tumor bed (4). Axillary lymph node response was evaluated according to morphologic and enhancement characteristics, including cortical thickness, perinodal infiltration, preservation or loss of the fatty hilum, and interval size change (18). Post-therapy breast lesions were classified as positive or negative for residual disease. For nodal visual analysis, the unit of analysis was the axillary compartment rather than the individual lymph node. Only baseline tracer-avid compartments were reassessed after treatment; a compartment was considered negative if no lymph node showed abnormal tracer uptake and positive if at least 1 lymph node remained tracer-avid.

### Statistical Analysis

Statistical analyses were performed using R (version 4.5.2) and IBM SPSS Statistics (version 26.0). Continuous variables were compared using the Student t test, Welch t test, or Mann-Whitney U test, as appropriate according to distributional characteristics and variance equality; categorical variables were analyzed using the χ² test or Fisher-Freeman-Halton exact test. Variables with statistical significance in univariable analyses were further explored using least absolute shrinkage and selection operator (LASSO) regression. Receiver-operating-characteristic (ROC) curves were constructed to evaluate diagnostic performance. The area under the curve (AUC), 95% confidence interval, sensitivity, specificity, and accuracy were calculated, and optimal cutoff values were determined by maximizing the Youden index. AUCs were compared using the DeLong test. To account for multiple comparisons in exploratory univariable analyses, false discovery rate was controlled using the Benjamini-Hochberg procedure, applied to families of imaging-parameter comparisons for primary breast lesions and axillary lymph nodes. A 2-sided P value less than 0.05 was considered statistically significant.

## RESULTS

### Patient Characteristics

Of the 27 enrolled patients, 24 completed the study protocol after exclusion of 3 patients (Fig. 2). Baseline characteristics are summarized in Table 1. All patients had AJCC stage II or III disease, including 1 case of occult breast cancer and 5 patients without axillary nodal metastasis at baseline. Because 1 patient had occult breast cancer without an identifiable breast lesion and 2 patients had 2 distinct breast lesions, lesion-level analysis included 25 primary breast lesions. Before NAC, imaging identified 44 metastatic lymph nodes across 27 axillary compartments. All tumors were invasive ductal carcinoma.

**FIGURE 2.**
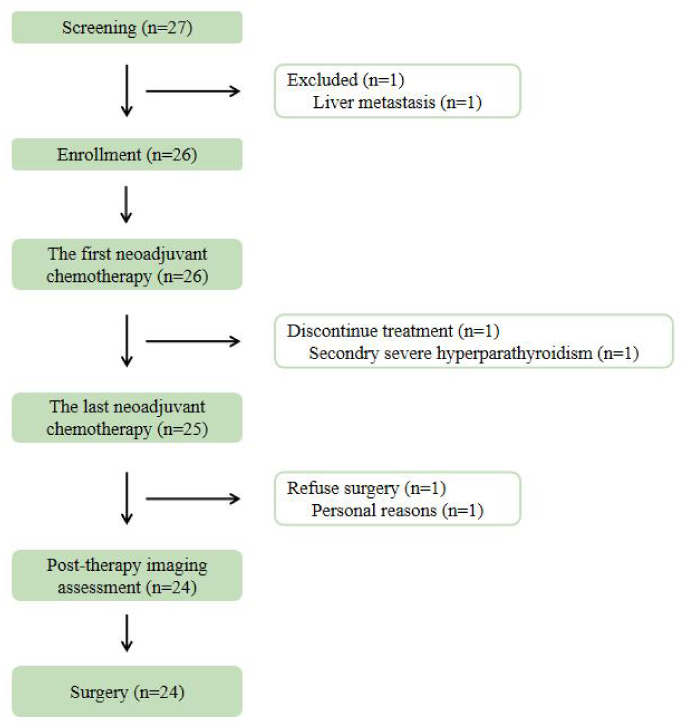
Trial profile.

**TABLE 1.** Patient Characteristics (n = 24)

| Characteristic | No. of patients |
| --- | --- |
| Mean age ± SD (y) | 49.29 ± 9.35(range, 36-64) |
| Mean BMI ± SD | 23.96 ± 2.08(range, 19.33-27.75) |
| Menstrual state |  |
| Postmenopause | 9(37.50%) |
| Premenopause | 15(62.50%) |
| Clinical T category |  |
| cT0 | 1(4.17%) |
| cT1 | 2(8.33%) |
| cT2 | 17(70.83%) |
| cT3 | 4(16.67%) |
| Clinical N category |  |
| cN0 | 5(20.83%) |
| cN1 | 13(54.17%) |
| cN2 | 2(8.33%) |
| cN3 | 4(16.67%) |
| AJCC clinical stage |  |
| Stage II (cT<sub>0-1</sub>N<sub>1</sub>M<sub>0</sub>/ cT<sub>2</sub>N<sub>0-1</sub>M<sub>0</sub>/ cT<sub>3</sub>N<sub>0</sub>M<sub>0</sub>) | 17(70.83%) |
| Stage III (cT<sub>3</sub>N<sub>1</sub>M<sub>0</sub>/ cT<sub>4</sub>N<sub>0-1</sub>M<sub>0</sub>/ cT<sub>0-4</sub>N<sub>2-3</sub>M<sub>0</sub>) | 7(29.17%) |
| Ki-67 |  |
| High expression | 13(54.17%) |
| Low expression | 11(45.83%) |
| Histological grading |  |
| G2 | 7(29.17%) |
| G3 | 17(70.83%) |
| Tumor type |  |
| Invasive ductal carcinoma | 24(100.00%) |
| Invasive lobular carcinoma | 0(0.00%) |
| Molecular subtype |  |
| Luminal | 4(16.67%) |
| HER2 positive | 12(50.00%) |
| Triple-negative | 8(33.33%) |
| Pathologic response (all lesions, n = 24) |  |
| pCR | 13(54.17%) |
| Non-pCR | 11(45.83%) |
| Pathologic response (breast primary lesions, n = 25) |  |
| pCR | 15(60.00%) |
| Non-pCR | 10(40.00%) |
| Pathologic response (axillary lymph nodes, n = 44) |  |
| pCR | 32(72.73%) |
| Non-pCR | 12(27.27%) |

HER2 = Human Epidermal Growth Factor Receptor 2; pCR = pathologic complete response. At lesion level, 25 primary breast lesions were analyzed because 1 patient had occult breast cancer without an identifiable breast lesion and 2 patients had 2 distinct primary lesions.

### Pathology Assessment

Among the 24 patients, 16 (66.67%) underwent modified radical mastectomy and 8 (33.33%) underwent breast-conserving surgery; 19 (79.17%) underwent axillary lymph node dissection and 5 (20.83%) underwent sentinel lymph node biopsy alone. The overall patient-level pCR rate was 54.17% (13/24). The lesion-level pCR rate for primary breast lesions was 60.00% (15/25), and the node-level pCR rate for axillary lymph nodes was 72.73% (32/44). Of the 15 primary lesions achieving pCR, 11 (73.33%) occurred in patients with AJCC stage II disease and 4 (26.67%) in patients with stage III disease. Among axillary lymph nodes achieving pCR, stage II and III disease each accounted for 50.00% (16/32).

Among the 10 patients who did not achieve pCR in primary lesions, 8 were classified as ypT1, 1 as ypT2, and 1 as ypT4 due to skin involvement. Among the 6 patients without nodal pCR, 4 were classified as ypN1, 1 as ypN2, and 1 as ypN3 due to supraclavicular nodal metastasis.

### Relationship Between Clinicopathologic Characteristics and pCR

Associations between clinicopathologic variables and pathologic response after NAC are summarized in Table 2. No clinicopathologic factor was significantly associated with pCR in primary tumors. For axillary nodal response, the post-therapy neutrophil-to-lymphocyte ratio (NLR) was significantly lower in the pCR group compared to the non-pCR group (median: 2.17 vs. 2.72; P = 0.03). No significant association was observed for overall lesion pCR (Supplemental Table 1).

**TABLE 2.**
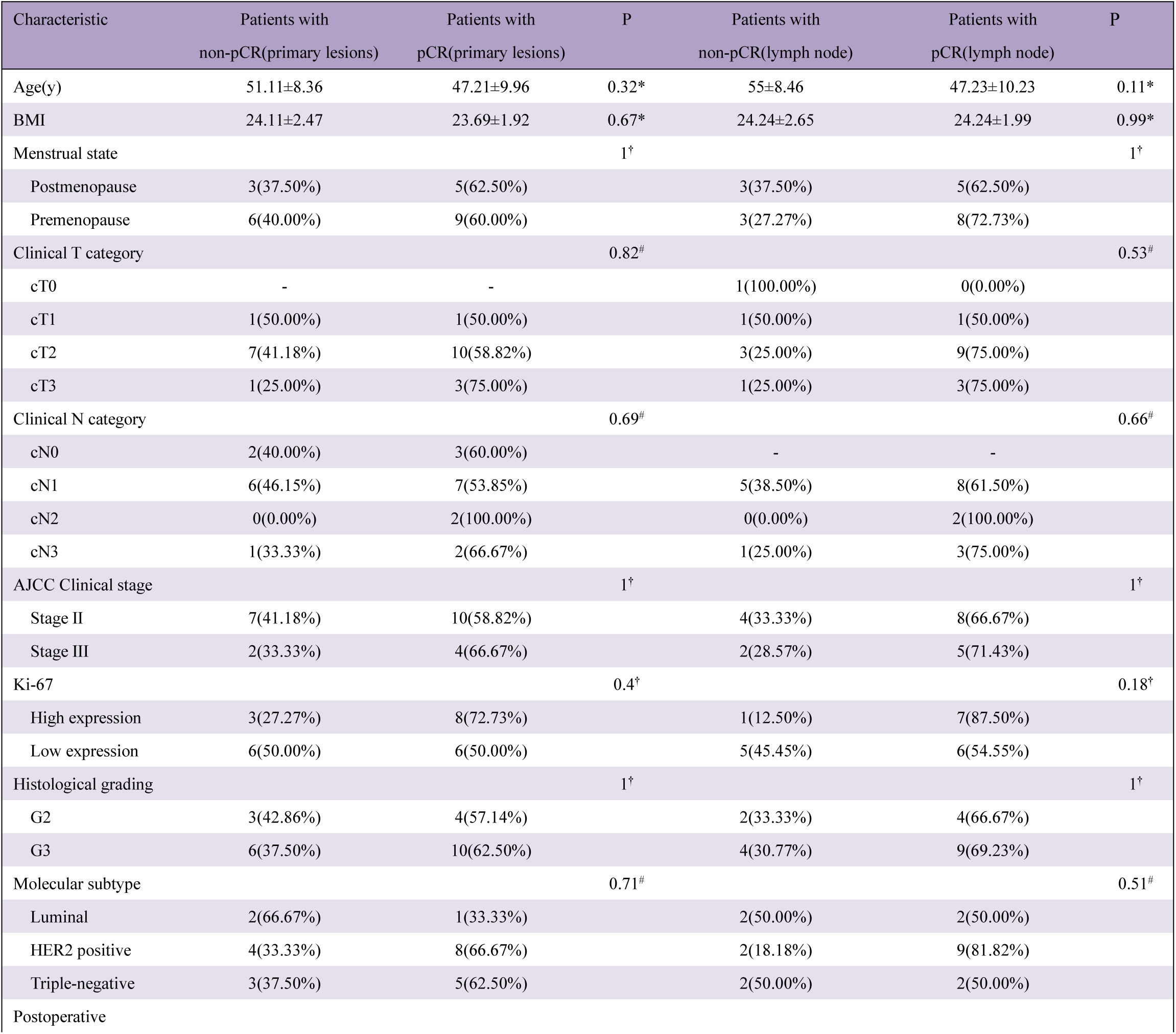
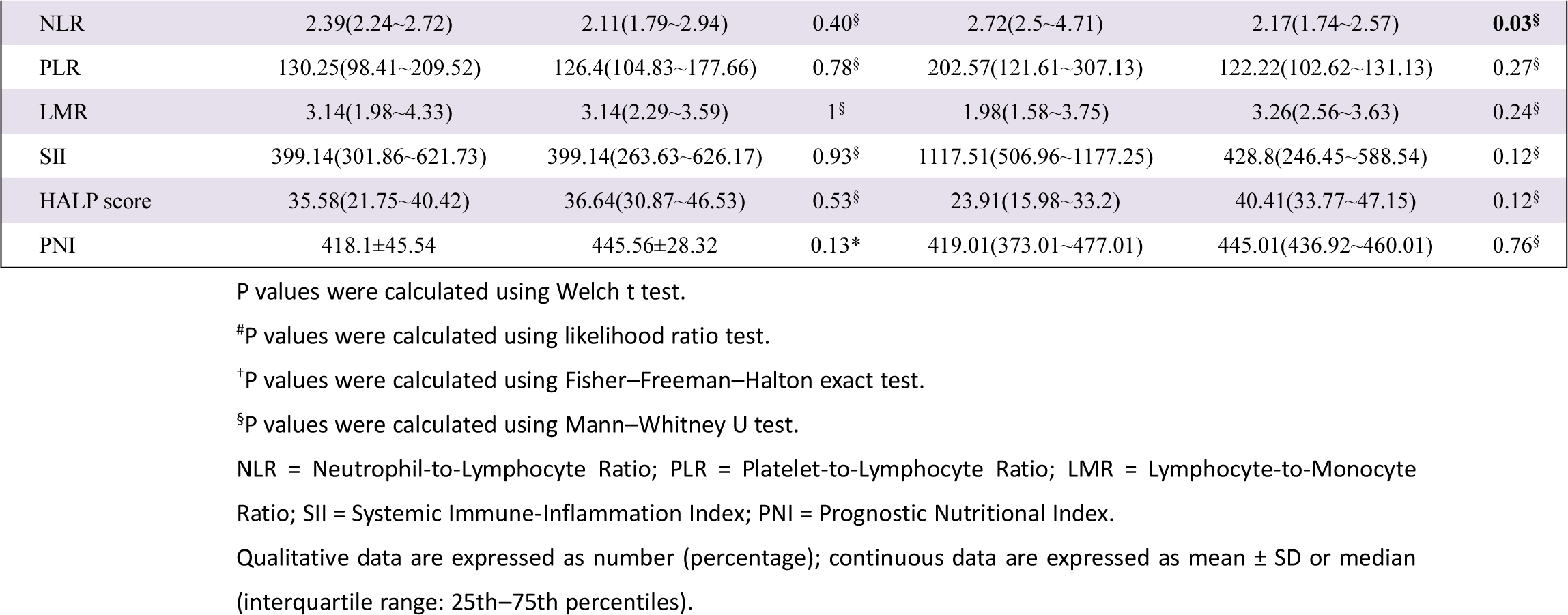
Clinicopathologic Characteristics of Patients According to Primary Lesions and Lymph Nodes Pathologic Response.

| Characteristic | Patients with non-pCR(primary lesions) | Patients with pCR(primary lesions) | P | Patients with non-pCR(lymph node) | Patients with pCR(lymph node) | P |
| --- | --- | --- | --- | --- | --- | --- |
| Age(y) | 51.11±8.36 | 47.21±9.96 | 0.32* | 55±8.46 | 47.23±10.23 | 0.11* |
| BMI | 24.11±2.47 | 23.69±1.92 | 0.67* | 24.24±2.65 | 24.24±1.99 | 0.99* |
| Menstrual state |  |  | 1<sup>†</sup> |  |  | 1<sup>†</sup> |
| Postmenopause | 3(37.50%) | 5(62.50%) |  | 3(37.50%) | 5(62.50%) |  |
| Premenopause | 6(40.00%) | 9(60.00%) |  | 3(27.27%) | 8(72.73%) |  |
| Clinical T category |  |  | 0.82<sup>#</sup> |  |  | 0.53<sup>#</sup> |
| cT0 | - | - |  | 1(100.00%) | 0(0.00%) |  |
| cT1 | 1(50.00%) | 1(50.00%) |  | 1(50.00%) | 1(50.00%) |  |
| cT2 | 7(41.18%) | 10(58.82%) |  | 3(25.00%) | 9(75.00%) |  |
| cT3 | 1(25.00%) | 3(75.00%) |  | 1(25.00%) | 3(75.00%) |  |
| Clinical N category |  |  | 0.69<sup>#</sup> |  |  | 0.66<sup>#</sup> |
| cN0 | 2(40.00%) | 3(60.00%) |  | - | - |  |
| cN1 | 6(46.15%) | 7(53.85%) |  | 5(38.50%) | 8(61.50%) |  |
| cN2 | 0(0.00%) | 2(100.00%) |  | 0(0.00%) | 2(100.00%) |  |
| cN3 | 1(33.33%) | 2(66.67%) |  | 1(25.00%) | 3(75.00%) |  |
| AJCC Clinical stage |  |  | 1<sup>†</sup> |  |  | 1<sup>†</sup> |
| Stage II | 7(41.18%) | 10(58.82%) |  | 4(33.33%) | 8(66.67%) |  |
| Stage III | 2(33.33%) | 4(66.67%) |  | 2(28.57%) | 5(71.43%) |  |
| Ki-67 |  |  | 0.4<sup>†</sup> |  |  | 0.18<sup>†</sup> |
| High expression | 3(27.27%) | 8(72.73%) |  | 1(12.50%) | 7(87.50%) |  |
| Low expression | 6(50.00%) | 6(50.00%) |  | 5(45.45%) | 6(54.55%) |  |
| Histological grading |  |  | 1<sup>†</sup> |  |  | 1<sup>†</sup> |
| G2 | 3(42.86%) | 4(57.14%) |  | 2(33.33%) | 4(66.67%) |  |
| G3 | 6(37.50%) | 10(62.50%) |  | 4(30.77%) | 9(69.23%) |  |
| Molecular subtype |  |  | 0.71<sup>#</sup> |  |  | 0.51<sup>#</sup> |
| Luminal | 2(66.67%) | 1(33.33%) |  | 2(50.00%) | 2(50.00%) |  |
| HER2 positive | 4(33.33%) | 8(66.67%) |  | 2(18.18%) | 9(81.82%) |  |
| Triple-negative | 3(37.50%) | 5(62.50%) |  | 2(50.00%) | 2(50.00%) |  |
| Postoperative |  |  |  |  |  |  |
| NLR | 2.39(2.24~2.72) | 2.11(1.79~2.94) | 0.40<sup>§</sup> | 2.72(2.5~4.71) | 2.17(1.74~2.57) | **0.03<sup>§</sup>** |
| PLR | 130.25(98.41~209.52) | 126.4(104.83~177.66) | 0.78<sup>§</sup> | 202.57(121.61~307.13) | 122.22(102.62~131.13) | 0.27<sup>§</sup> |
| LMR | 3.14(1.98~4.33) | 3.14(2.29~3.59) | 1<sup>§</sup> | 1.98(1.58~3.75) | 3.26(2.56~3.63) | 0.24<sup>§</sup> |
| SII | 399.14(301.86~621.73) | 399.14(263.63~626.17) | 0.93<sup>§</sup> | 1117.51(506.96~1177.25) | 428.8(246.45~588.54) | 0.12<sup>§</sup> |
| HALP score | 35.58(21.75~40.42) | 36.64(30.87~46.53) | 0.53<sup>§</sup> | 23.91(15.98~33.2) | 40.41(33.77~47.15) | 0.12<sup>§</sup> |
| PNI | 418.1±45.54 | 445.56±28.32 | 0.13* | 419.01(373.01~477.01) | 445.01(436.92~460.01) | 0.76<sup>§</sup> |

P values were calculated using Welch t test.
<sup>#</sup>P values were calculated using likelihood ratio test.
<sup>†</sup>P values were calculated using Fisher–Freeman–Halton exact test.
<sup>§</sup>P values were calculated using Mann–Whitney U test.
NLR = Neutrophil-to-Lymphocyte Ratio; PLR = Platelet-to-Lymphocyte Ratio; LMR = Lymphocyte-to-Monocyte Ratio; SII = Systemic Immune-Inflammation Index; PNI = Prognostic Nutritional Index.
Qualitative data are expressed as number (percentage); continuous data are expressed as mean ± SD or median (interquartile range: 25th–75th percentiles).

**SUPPLEMENTAL TABLE 1.**
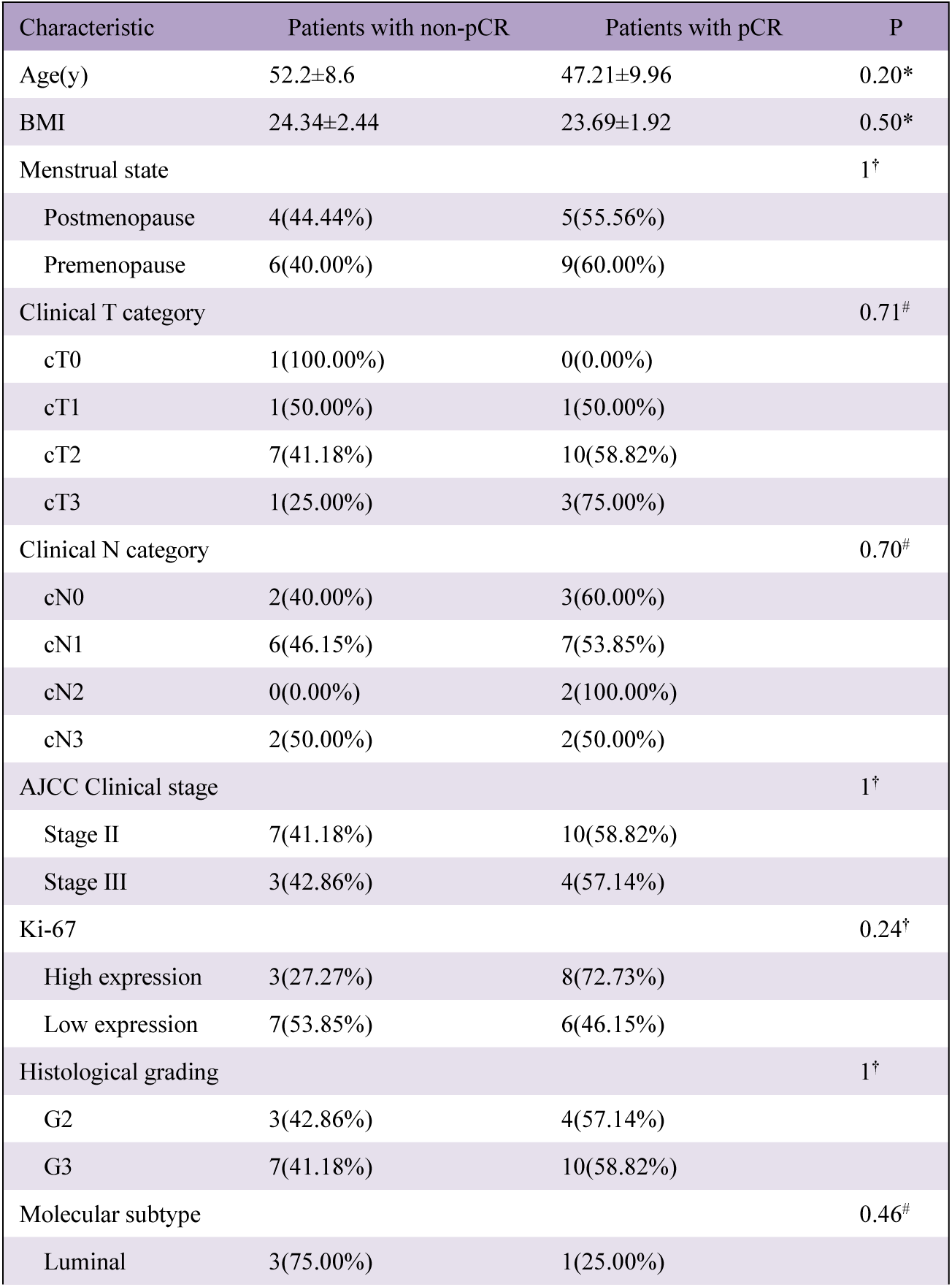
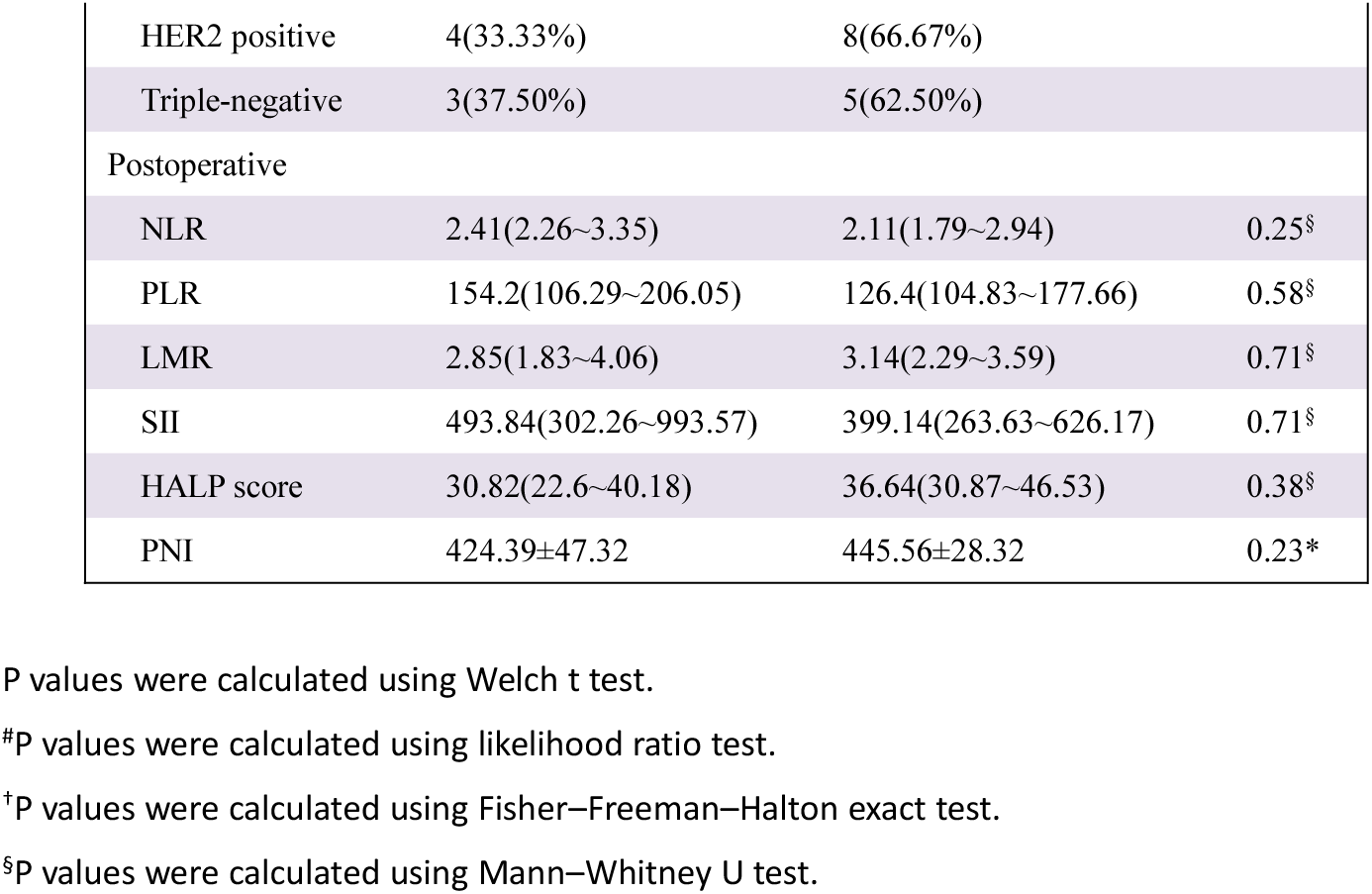
Clinical Characteristics of Patients According to All Lesions Pathologic Response.

### Visual Assessment of [^68^Ga]Ga-FAPI-04 and [^18^F]FDG in post-therapy

Post-therapy PET images were visually assessed for prediction of pathologic response. Axillary lymph nodes were evaluated according to levels I-III, and only baseline tracer-avid compartments were included. A compartment was defined as negative if no lymph node exhibited abnormal tracer uptake and positive if at least 1 lymph node demonstrated tracer avidity. For primary lesions (Table 3), visual assessment using [^68^Ga]Ga-FAPI-04 showed higher accuracy than [^18^F]FDG, although the difference was not statistically significant. For compartment-level nodal pCR (Table 4), [^68^Ga]Ga-FAPI-04 showed significantly higher diagnostic performance than [^18^F]FDG and was significantly associated with nodal pCR (P < 0.01), with a sensitivity of 87.50%, specificity of 84.21%, and accuracy of 85.19%. Representative cases are shown in Figure 3.

**TABLE 3.**
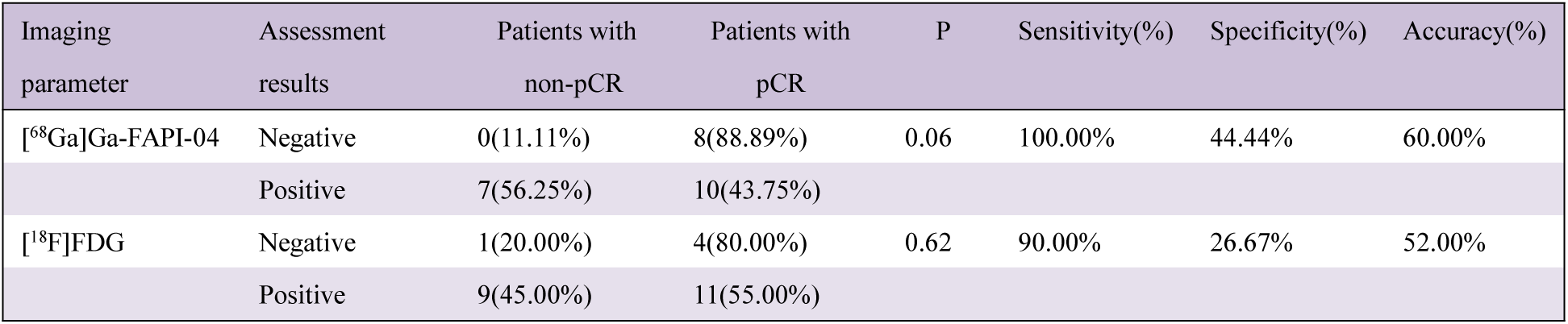
Visual Assessment of [^68^Ga]Ga-FAPI-04 and [^18^F]FDG for Primary Lesion pCR in post-therapy.

| Imaging parameter | Assessment results | Patients with non-pCR | Patients with pCR | P | Sensitivity(%) | Specificity(%) | Accuracy(%) |
| --- | --- | --- | --- | --- | --- | --- | --- |
| [<sup>68</sup>Ga]Ga-FAPI-04 | Negative | 0(11.11%) | 8(88.89%) | 0.06 | 100.00% | 44.44% | 60.00% |
|  | Positive | 7(56.25%) | 10(43.75%) |  |  |  |  |
| [<sup>18</sup>F]FDG | Negative | 1(20.00%) | 4(80.00%) | 0.62 | 90.00% | 26.67% | 52.00% |
|  | Positive | 9(45.00%) | 11(55.00%) |  |  |  |  |

**TABLE 4.**
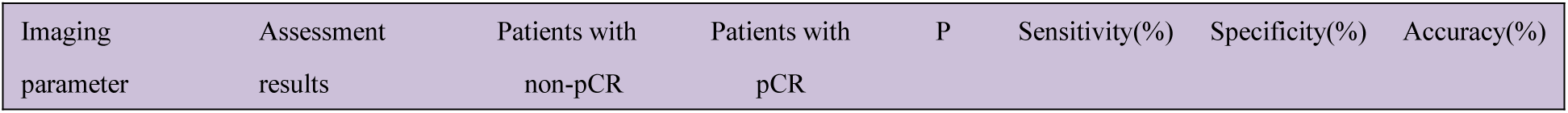
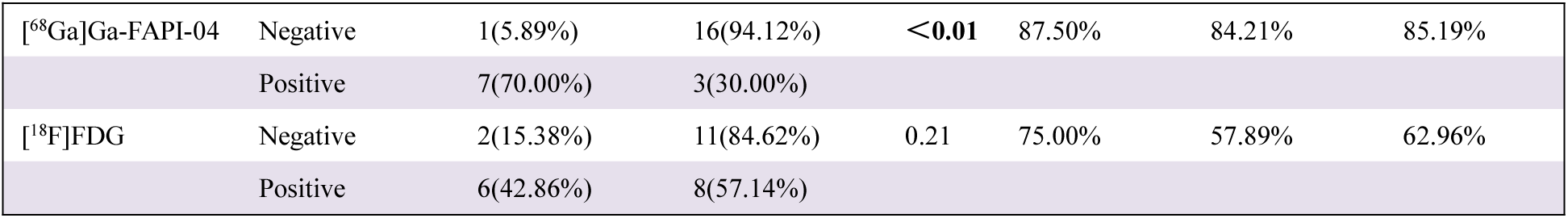
Visual Assessment of [^68^Ga]Ga-FAPI-04 and [^18^F]FDG for Axillary Compartment-Level Nodal pCR in post-therapy.

**FIGURE 3.**
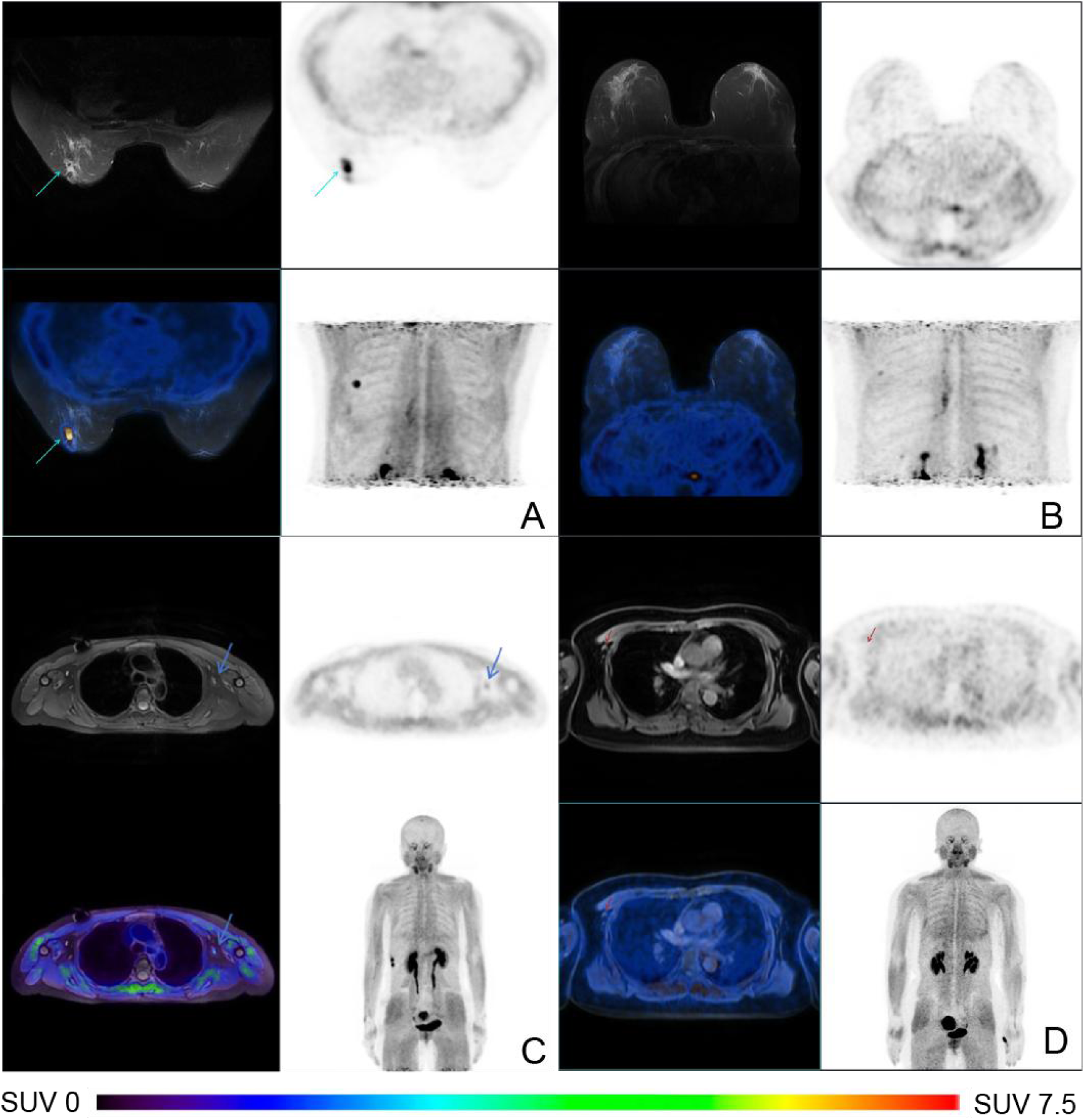
Post-therapy [^68^Ga]Ga-FAPI-04 PET/MRI images.A Visual assessment positive for the primary lesion; B visual assessment negative for the primary lesion; C visual assessment positive for axillary lymph nodes; D visual assessment negative for axillary lymph nodes.

### Performance of [^68^Ga]Ga-FAPI-04 PET/MRI, [^18^F]FDG PET/CT, and Contrast-Enhanced MRI

In quantitative analyses (Table 5), no baseline [^68^Ga]Ga-FAPI-04 or [^18^F]FDG PET parameter was significantly associated with primary-lesion pCR. On post-therapy [^68^Ga]Ga-FAPI-04 imaging, SUVmax, SUVmean, SULmax, SULmean, SULpeak, and TBR were significantly lower in the pCR group (all P < 0.05), whereas no post-therapy [^18^F]FDG PET parameter showed a significant association. SUVmax, SULmax, SULmean and TBR also remained significant after false discovery rate correction, whereas SUVmean and SULpeak did not. For percentage changes, ΔFTV% and ΔTLF% on [^68^Ga]Ga-FAPI-04 PET, as well as ΔSULmax%, ΔSULpeak%, and ΔTBR% on [^18^F]FDG PET, were significantly higher in the pCR group before correction; however, after false discovery rate correction, only ΔSULpeak% and ΔTBR% remained statistically significant. (Supplemental Table 8).

**TABLE 5.** PET Characteristics According to Pathologic Response in Primary Lesions and Axillary Lymph Nodes.

| Imaging parameter | Subparameter | Patients with non-pCR(primary lesions) | Patients with pCR(primary lesions) | P | Patients with non-pCR(lymph node) | Patients with pCR(lymph node) | P |
| --- | --- | --- | --- | --- | --- | --- | --- |
| Pre-therapy |  |  |  |  |  |  |  |
| [$^{68}\text{Ga}$]Ga-FAPI-04 | SUVmax | 17.18±6.64 | 15.26±6.7 | 0.49* | 9.79(8.13~12.25) | 7.92(4.56~10.08) | 0.08$^{\S}$ |
| | SUVmean | 9.31±4.78 | 7.37±3.23 | 0.28* | 6.42(4.94~7.03) | 4.89(2.46~5.96) | 0.06$^{\S}$ |
| | SULmax | 12.27±4.59 | 11.05±5.03 | 0.54* | 6.81(5.7~8.59) | 5.3(3.36~7.22) | 0.8$^{\S}$ |
| | SULmean | 6.64±3.32 | 5.34±2.44 | 0.31* | 4.15±1.38 | 3.31±1.79 | 0.11* |
| | SULpeak | 9.29±4.19 | 8.62±4.29 | 0.70* | 4.51(3.71~6.5) | 3.36(1.9~5.15) | 0.06$^{\S}$ |
| | FTV($\text{cm}^3$) | 16.87±12.43 | 18.74±15.47 | 0.74* | 4.62±5.82 | 3.95±10.19 | 0.79* |
| | TLF (SUV$\cdot\text{cm}^3$) | 93.34(60.19~174.32) | 112.99(47.51~244.94) | 1$^{\S}$ | 32.12±49.09 | 19.66±48.36 | 0.46* |
| | TBR | 11.39±6.25 | 14.36±11.56 | 0.41* | 12.13±5.23 | 7.84±5.23 | **0.03\*** |
| [$^{18}\text{F}$]FDG | SUVmax | 8.27(6.44~10.26) | 12.73(6.76~15.76) | 0.40$^{\S}$ | 5.12(3.1~9.74) | 6.05(4.98~10.01) | 0.59$^{\S}$ |
| | SUVmean | 3.63(3.42~4.11) | 5.21(3.67~7.85) | 0.26$^{\S}$ | 2.82(2.32~4.16) | 3.24(2.5~4.94) | 0.67$^{\S}$ |
| | SULmax | 6.8±2.95 | 8.86±5.04 | 0.21* | 3.62(2.39~6.82) | 4.26(3.48~7.23) | 0.56$^{\S}$ |
| | SULmean | 2.55(2.49~2.96) | 3.65(2.68~5.66) | 0.26$^{\S}$ | 1.98(1.64~2.92) | 2.37(1.76~3.61) | 0.62$^{\S}$ |
| | SULpeak | 3.89(3.46~4.29) | 6.1(4~9.04) | 0.24$^{\S}$ | 2(1.64~3.59) | 2.29(1.67~4.04) | 0.97$^{\S}$ |
| | MTV($\text{cm}^3$) | 14.02±10.53 | 12.8±10.3 | 0.78* | 1.65(0.47~6.2) | 1.23(0.76~1.7) | 0.95$^{\S}$ |
| | TLG (SUV$\cdot\text{cm}^3$) | 45.04(25.92~78.38) | 50.66(24.34~117.83) | 0.85$^{\S}$ | 5.16(0.98~21.25) | 3.68(1.83~7.95) | 0.84$^{\S}$ |
| | TBR | 5.9(4.18~7.86) | 6.94(4.33~14.15) | 0.46$^{\S}$ | 6.88(5.2~9.69) | 4.91(3.51~7.51) | 0.10$^{\S}$ |
| Post-therapy |  |  |  |  |  |  |  |
| [$^{68}\text{Ga}$]Ga-FAPI-04 | SUVmax | 2.73(2.41~4.02) | 1.04(0.72~2.24) | **<0.01$^{\S}$** | 2.09(0.86~5.27) | 0.58(0.52~1.07) | **<0.01$^{\S}$** |
|  | SUVmean | 1.62(1.45~2.62) | 0.82(0.48~1.52) | **0.02$^{\S}$** | 1.6(0.53~3.16) | 0.41(0.36~0.44) | **<0.01$^{\S}$** |
|  | SULmax | 1.94(1.68~3.04) | 0.82(0.54~1.85) | **0.01$^{\S}$** | 1.54(0.6~3.69) | 0.42(0.37~0.78) | **<0.01$^{\S}$** |
|  | SULmean | 1.16(1.03~1.97) | 0.58(0.36~1.1) | **<0.01$^{\S}$** | 1.17(0.37~2.21) | 0.3(0.25~0.33) | **<0.01$^{\S}$** |
|  | SULpeak | 1.27(1.05~1.72) | 0.58(0.36~1.23) | **0.02$^{\S}$** | 0.92(0.37~1.84) | 0.3(0.25~0.33) | **<0.01$^{\S}$** |
| | FTV($\text{cm}^3$) | 1.34(1.07~2.24) | 0(0~1.73) | 0.18$^{\S}$ | 0.62(0~0.8) | 0(0~0) | **<0.01$^{\S}$** |
| | TLF (SUV$\cdot\text{cm}^3$) | 2.19(1.75~6.42) | 0(0~2.66) | 0.15$^{\S}$ | 0.89(0~2.51) | 0(0~0) | **<0.01$^{\S}$** |
|  | TBR | 2.64(2.32~3.83) | 1.8(1~2.53) | **0.01$^{\S}$** | 2.88(0.8~5.17) | 0.96(0.55~1.02) | **0.01$^{\S}$** |
| [$^{18}\text{F}$]FDG | SUVmax | 1.58(1.53~1.95) | 1.58(1.25~1.77) | 0.27$^{\S}$ | 0.64(0.64~1.86) | 0.57(0.44~1.06) | 0.10$^{\S}$ |
| | SUVmean | 1.17(1.13~1.48) | 1.12(0.7~1.32) | 0.29$^{\S}$ | 0.34(0.34~0.9) | 0.32(0.26~0.71) | 0.23$^{\S}$ |
| | SULmax | 1.17(1.09~1.43) | 1.07(0.88~1.29) | 0.20$^{\S}$ | 0.45(0.45~1.4) | 0.42(0.34~0.71) | 0.16$^{\S}$ |
|  | SULmean | 0.88(0.82~1.07) | 0.85(0.5~0.94) | 0.21<sup>§</sup> | 0.24(0.24~0.65) | 0.22(0.19~0.46) | 0.13<sup>§</sup> |
|  | SULpeak | 0.86(0.84~1.06) | 0.82(0.52~0.96) | 0.23<sup>§</sup> | 0.24(0.24~0.64) | 0.22(0.19~0.4) | 0.12<sup>§</sup> |
|  | MTV(cm<sup>3</sup>) | 1.71(1.54~2.4) | 1.4(0.25~2.69) | 0.19<sup>§</sup> | 0(0~0.45) | 0(0~0.35) | 0.89<sup>§</sup> |
|  | TLG (SUV·cm<sup>3</sup>) | 2.45(1.83~3.99) | 1.75(0.3~2.74) | 0.16<sup>§</sup> | 0(0~0.55) | 0(0~0.31) | 0.76<sup>§</sup> |
|  | TBR | 2.98±1.49 | 2.43±1.21 | 0.34* | 1(0.96~2.99) | 0.84(0.68~1.41) | 0.11<sup>§</sup> |
| MRI result |  |  |  | **0.02<sup>†</sup>** |  |  | **0.01<sup>†</sup>** |
| Negative |  | 2(15.38%) | 11(84.62%) |  | 2(11.76%) | 15(88.24%) |  |
| Positive |  | 8(66.67%) | 4(33.33%) |  | 6(60.00%) | 4(40.00%) |  |
| Pre- to post-therapy percentage change |  |  |  |  |  |  |  |
| <sup>68</sup>Ga]Ga-FAPI-04 | Δ SUVmax% | 80.85(77.82~86.84) | 87.95(81.41~94.89) | 0.10<sup>§</sup> | 79.55(47.31~88.32) | 88.69(79.52~94.12) | **0.02<sup>§</sup>** |
|  | Δ SUVmean% | 74.49(67.9~85.86) | 85.49(75.9~92.26) | 0.22<sup>§</sup> | 78.14(52.4~81.13) | 89.75(81.72~93) | **<0.01<sup>§</sup>** |
|  | Δ SULmax% | 89.02(84.3~91.92) | 93.3(88.88~96.25) | 0.13<sup>§</sup> | 79.14(47.37~88.15) | 88.72(79.7~94.16) | **0.03<sup>§</sup>** |
|  | Δ SULmean% | 75.69(65.49~85.93) | 85.48(76.46~92.47) | 0.20<sup>§</sup> | 78.69(52.48~80.93) | 89.76(81.79~93.14) | **<0.01<sup>§</sup>** |
|  | Δ SULpeak% | 84.74(74.82~90.31) | 91.17(83.09~95.31) | 0.16<sup>§</sup> | 82.17(62.25~86.63) | 89.49(83.38~93.78) | **0.03<sup>§</sup>** |
|  | Δ FTV% | 82.65(80.73~91.75) | 100(88.44~100) | **0.02<sup>§</sup>** | 85.64(65.43~100) | 100(100~100) | **<0.01<sup>§</sup>** |
|  | Δ TLF% | 97.28(91.01~98.8) | 100(96.69~100) | **0.03<sup>§</sup>** | 96.31(80.22~100) | 100(100~100) | **<0.01<sup>§</sup>** |
|  | Δ TBR% | 66.78(60.44~79.44) | 76.42(62.76~93.6) | 0.31<sup>§</sup> | 79.57(66.13~85.77) | 87.7(79.79~92.12) | 0.13<sup>§</sup> |
| <sup>18</sup>F]FDG | Δ SUVmax% | 77.5(73.74~84.82) | 87.72(83.43~89.84) | 0.06<sup>§</sup> | 79.25(76.59~88.86) | 90.48(81.67~92.86) | 0.09<sup>§</sup> |
|  | Δ SUVmean% | 64.97±19.54 | 78.15±14.18 | 0.09* | 85.13(75.92~89.47) | 87.83(83.6~92.52) | 0.29<sup>§</sup> |
|  | Δ SULmax% | 77.85(72.46~84.37) | 87.97(83.62~90.08) | **0.04<sup>§</sup>** | 80.43(76.26~88.44) | 90.3(82.1~92.91) | 0.09<sup>§</sup> |
|  | Δ SULmean% | 64.8±18.96 | 78.23±14.12 | 0.07* | 84.98(77.25~89.4) | 88.12(83.7~92.54) | 0.29<sup>§</sup> |
|  | Δ SULpeak% | 77.59(74~80.13) | 88.19(81.77~90.15) | **<0.01<sup>§</sup>** | 84.59(77.96~92.34) | 88.65(84.66~92.97) | 0.27<sup>§</sup> |
|  | Δ MTV% | 81.52(74.37~93.39) | 86.38(66.19~99.18) | 0.76<sup>§</sup> | 100(95.22~100) | 100(88.25~100) | 0.99<sup>§</sup> |
|  | Δ TLG% | 95.42(90.59~97.72) | 97.28(93.6~99.78) | 0.28<sup>§</sup> | 100(98.5~100) | 100(98.87~100) | 0.94<sup>§</sup> |
|  | Δ TBR% | 52.36(37.27~69.13) | 74.41(69.68~79.14) | **0.01<sup>§</sup>** | 79.51(64.4~87.16) | 82.05(74.42~87.36) | 0.97<sup>§</sup> |

\*P values were calculated using Welch t test.
<sup>†</sup>P values were calculated using Fisher–Freeman–Halton exact test.
<sup>§</sup>P values were calculated using Mann–Whitney U test.
Qualitative data are presented as number and percentage; continuous data are expressed as mean ± SD or median and 25th-75th percentiles. d indicates percentage change from pre-therapy to post-therapy.

For axillary lymph nodes, baseline [^68^Ga]Ga-FAPI-04 TBR showed a nominal association with nodal pCR (7.84 ± 5.23 vs. 12.13 ± 5.23; P = 0.03) but did not remain significant after false discovery rate correction. On post-therapy imaging, all evaluated [^68^Ga]Ga-FAPI-04 parameters remained significantly lower in the pCR group, and the post-therapy FAPI parameters remained significant after false discovery rate correction (all q ≤ 0.038). Among percentage-change metrics, Δ SUVmean%, Δ SULmean%, Δ FTV%, and Δ TLF% also remained significant after false discovery rate correction, whereas Δ SUVmax%, Δ SULmax%, and Δ SULpeak% did not (Supplemental Table 8).

Contrast-enhanced MRI detected all primary lesions and all axillary metastases at baseline. After treatment, 13 of 25 primary lesions were MRI-negative and 12 were MRI-positive; pCR was observed in 11 MRI-negative lesions, whereas 8 MRI-positive lesions exhibited residual disease (P < 0.05). Among 27 axillary compartments, 17 were MRI-negative and 10 were MRI-positive; nodal pCR was observed in 15 MRI-negative compartments, whereas residual metastatic lymph nodes were identified in 6 MRI-positive compartments (P < 0.05).

As shown in Supplemental Table 2, post-therapy [^68^Ga]Ga-FAPI-04 TLR showed a nominal association with overall-lesion pCR (P < 0.05), whereas no [^18^F]FDG parameter was associated with overall-lesion pCR. This association did not remain significant after false discovery rate correction (Supplemental Table 8).

**SUPPLEMENTAL TABLE 2.**
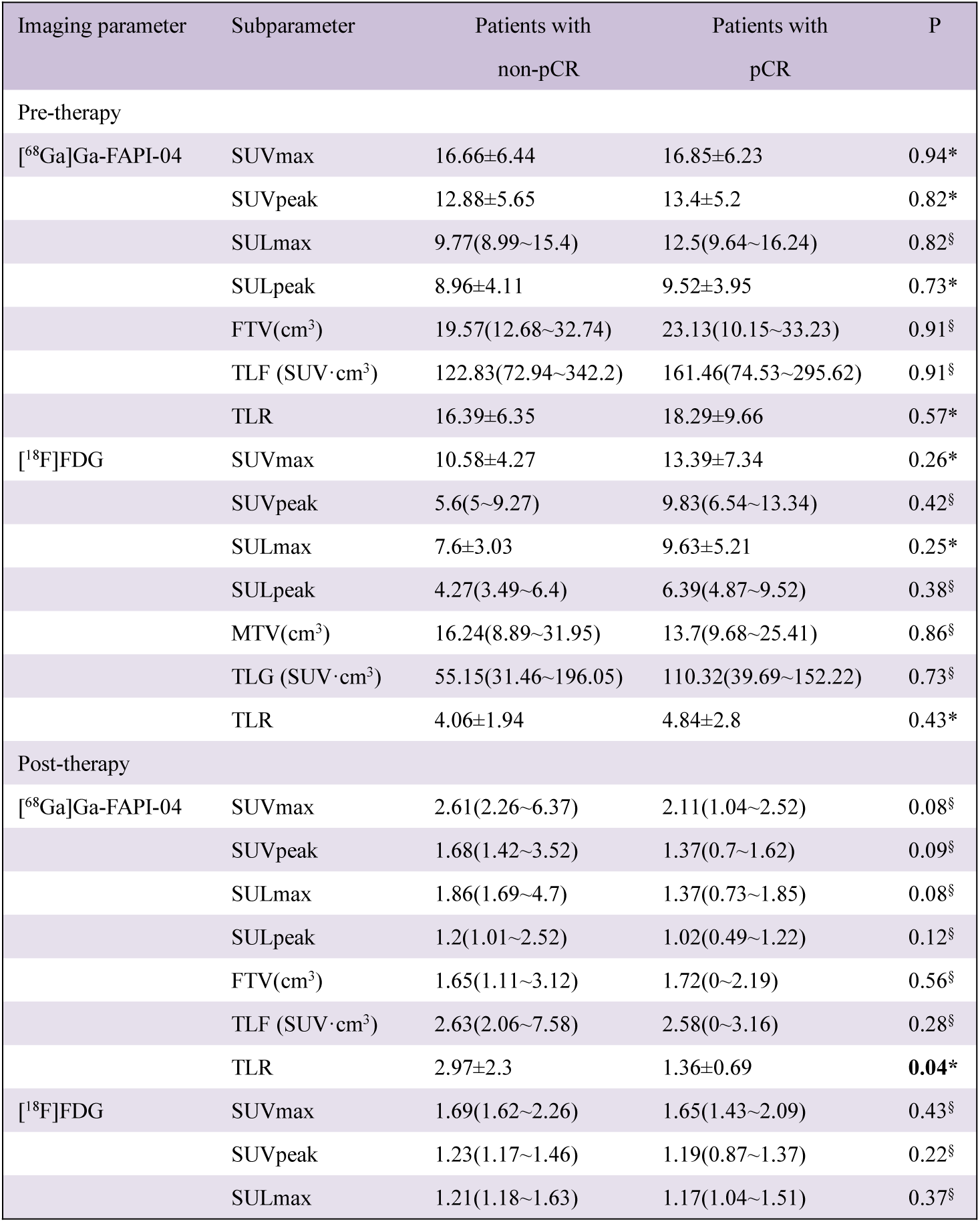
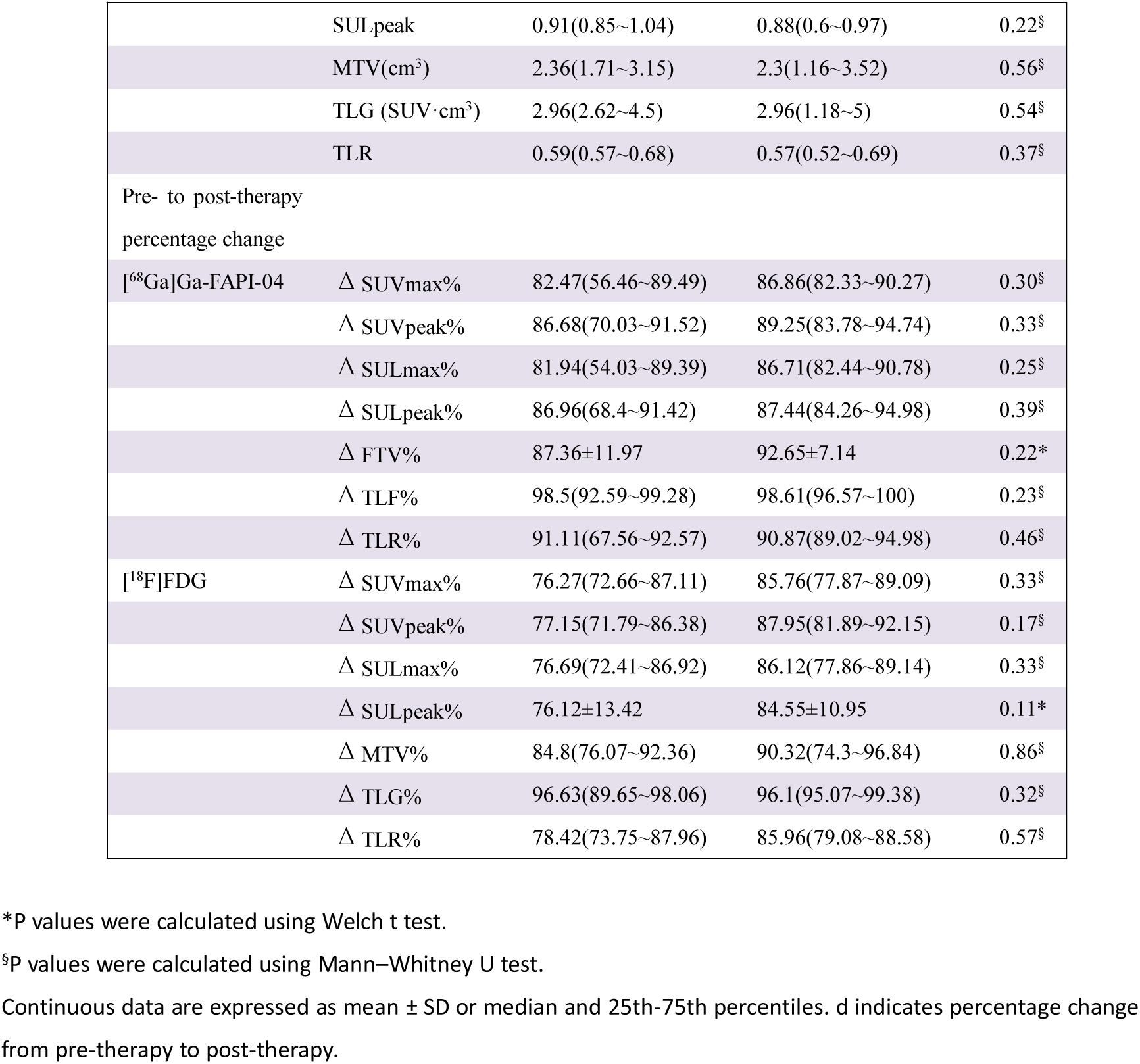
PET Characteristics of Patients According to Pathologic Response.

### Receiver-Operating-Characteristic Analyses of Imaging and Clinical Parameters for pCR

Variables were entered into LASSO regression for feature selection (Supplemental Figs. 1 and 2). For [^68^Ga]Ga-FAPI-04, post-therapy SUVmax, SULmean, and TBR were strongly associated with primary-lesion pCR, whereas post-therapy SULmean and Δ TLF% were associated with nodal pCR. For [^18^F]FDG, only Δ TBR% was associated with primary-lesion pCR; no significant parameter was identified for nodal pCR, and therefore LASSO analysis was not performed for nodal [^18^F]FDG parameters.

**SUPPLEMENTAL FIGURE 1.**
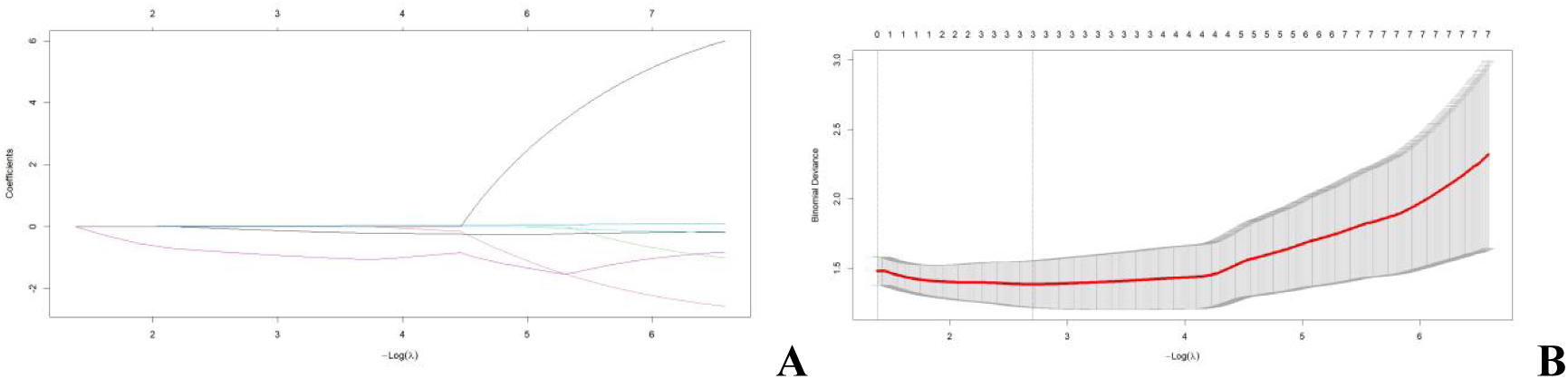
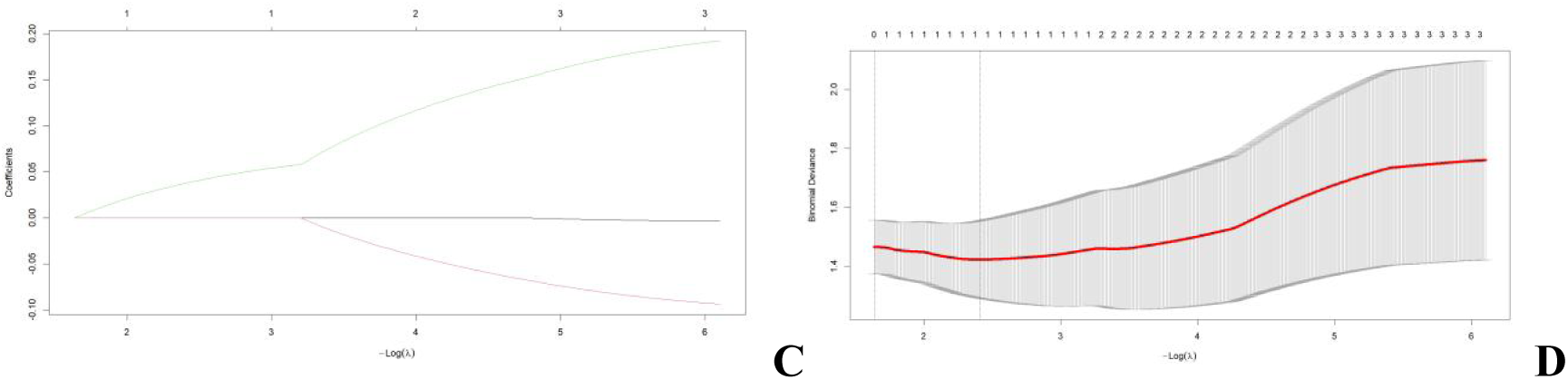
LASSO coefficient profiles for primary-lesion analyses. A and B show the coefficient profile plot and cross-validation curve for [68Ga]Ga-FAPI-04, respectively. C and D show the coefficient profile plot and cross-validation curve for [18F]FDG, respectively. The optimal lambda was selected on the basis of minimum binomial deviance.

**SUPPLEMENTAL FIGURE 2.**
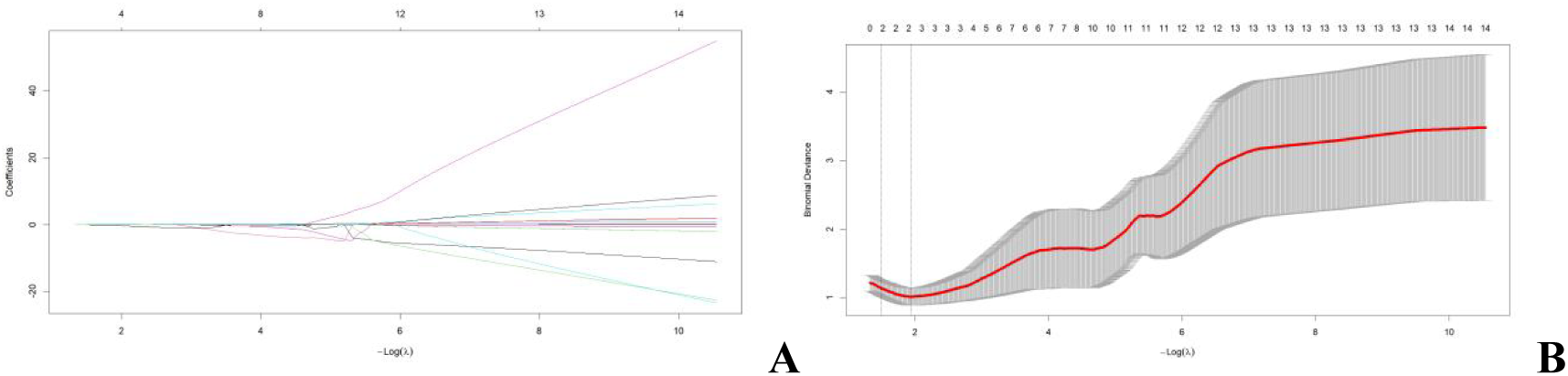
LASSO coefficient profiles for axillary nodal analyses. A shows the coefficient profile plot. B shows the cross-validation curve. The optimal lambda was selected on the basis of minimum binomial deviance.

ROC analysis identified the optimal parameters for predicting primary-lesion pCR (Fig. 4A, Table 6). Post-therapy [^68^Ga]Ga-FAPI-04 SUVmax showed the highest diagnostic performance (AUC, 0.84; 95% CI, 0.683-0.997; sensitivity, 80.00%; specificity, 80.00%; accuracy, 80.00%), whereas ΔTBR% was the best-performing [^18^F]FDG parameter (AUC, 0.747; 95% CI, 0.545-0.949; sensitivity, 70.00%; specificity, 80.00%; accuracy, 76.00%). DeLong testing showed no significant difference between the 2 tracers (P > 0.05). A representative case is shown in Figure 5.

**FIGURE 4.**
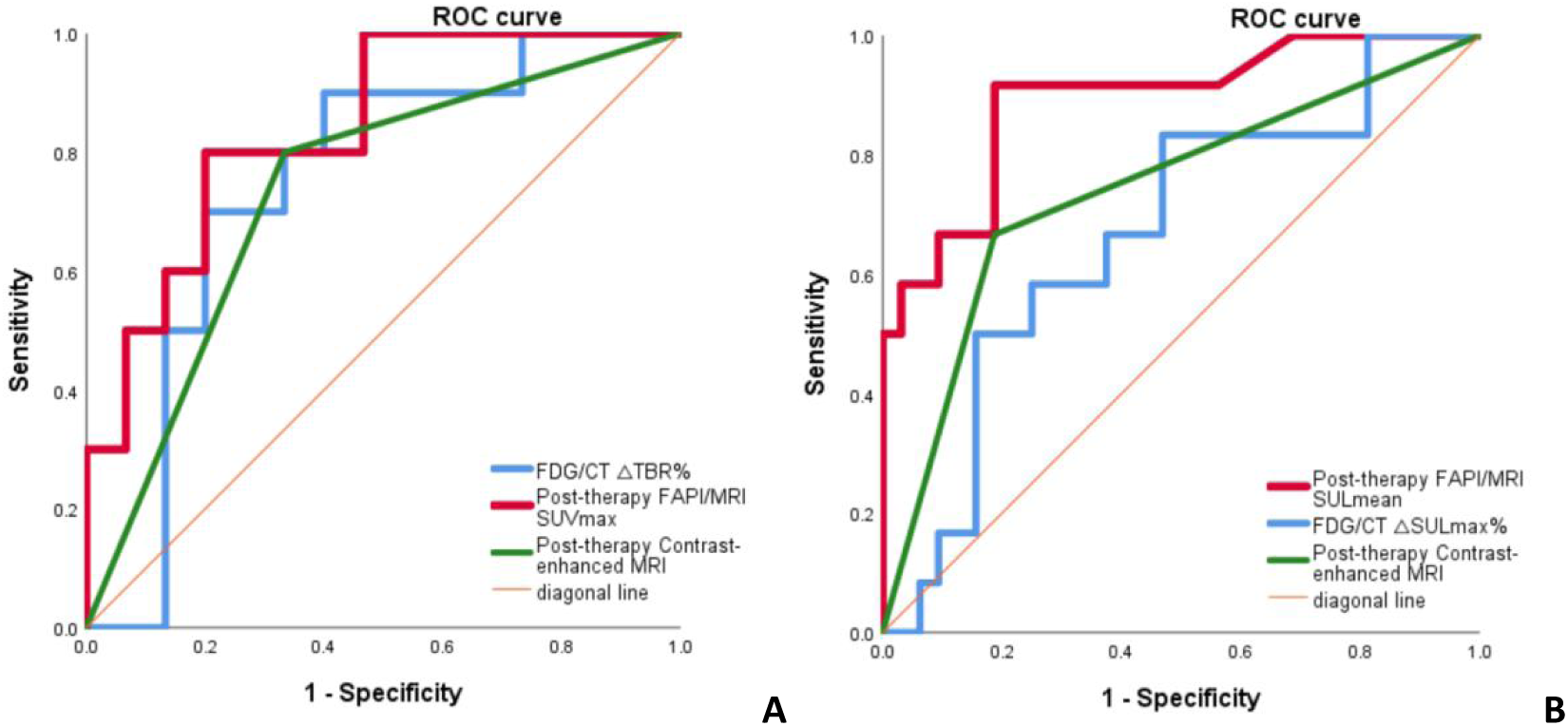
Receiver-operating-characteristic (ROC) curves of the optimal imaging parameters for prediction of pathologic complete response (pCR). A: ROC curves of post-therapy [^68^Ga]Ga-FAPI-04 SUVmax, [^18^F]FDG ΔTBR%, and contrast-enhanced MRI for predicting pCR in primary breast lesions. B: ROC curves of post-therapy [^68^Ga]Ga-FAPI-04 SULmean, [^18^F]FDG Δ SULmax%, and contrast-enhanced MRI for predicting pCR in axillary lymph nodes.

**TABLE 6.** Diagnostic Performance of Imaging Parameters for Prediction of pCR in Primary Lesions.

| Parameter | Subparameter | Cutoff | AUC | P | 95%CI | Sensitivity(%) | Specificity(%) | Accuracy(%) |
| --- | --- | --- | --- | --- | --- | --- | --- | --- |
| Contrast-enhanced MRI in post-therapy |  | — | 0.733 | **0.04** | 0.528-0.938 | 80.00% | 66.67% | 72.00% |
| $[^{68}\text{Ga}]\text{Ga-FAPI-04}$ | Post-therapy SUVmax | 2.395 | 0.84 | **<0.01** | 0.683-0.997 | 80.00% | 80.00% | 80.00% |
| $[^{18}\text{F}]\text{FDG}$ | $\Delta \text{TBR\%}$ | 59.05 | 0.747 | **0.04** | 0.545-0.949 | 70.00% | 80.00% | 76.00% |

AUC = Area Under the Curve.

**FIGURE 5.**
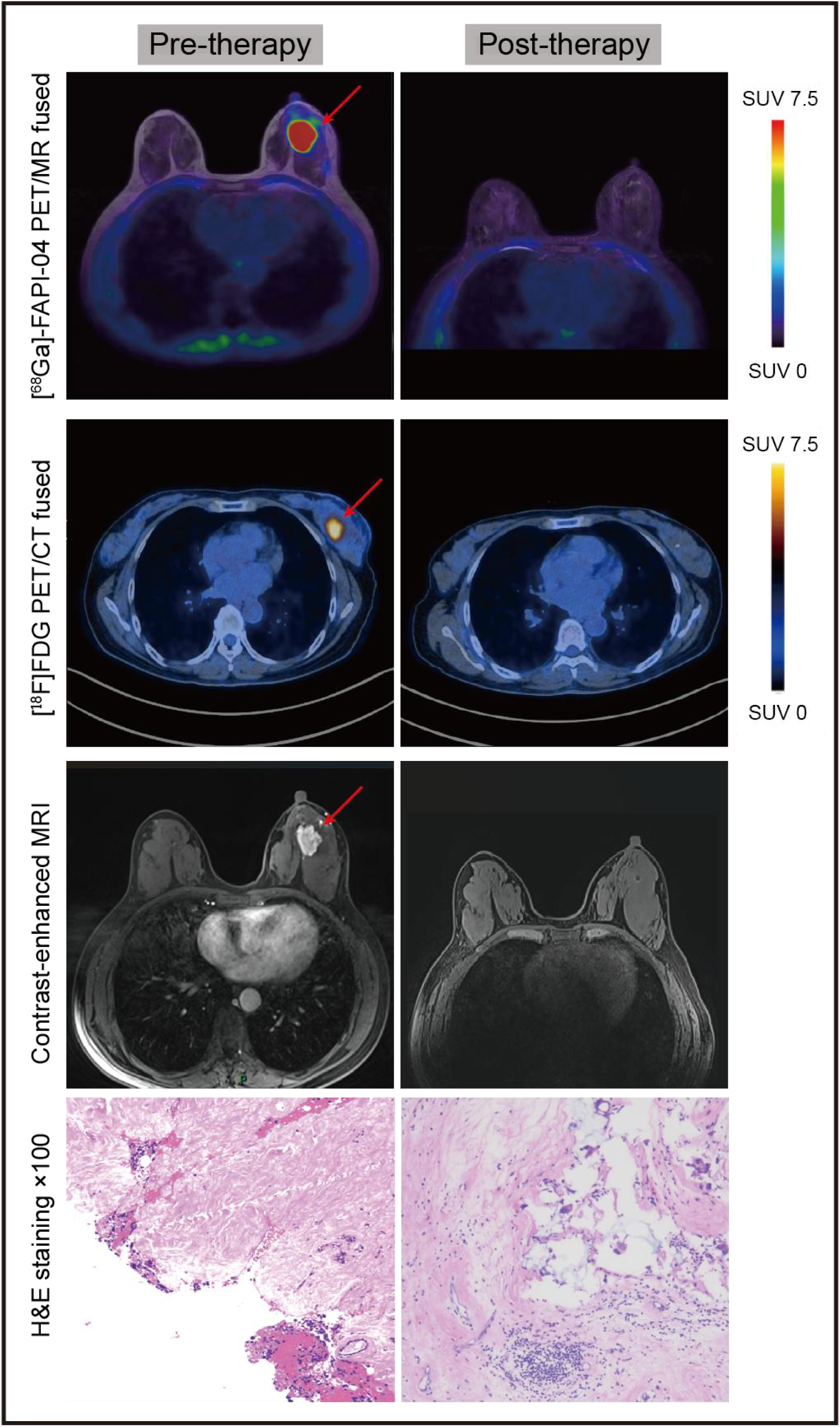
A woman with invasive breast cancer who achieved a pCR after NAC. Pre-therapy images obtained with three imaging modalities show the primary lesion (red arrows), which was confirmed as invasive breast carcinoma by core needle biopsy. Post-therapy images show no residual lesion on any modality (no significant tracer uptake on PET and no enhancement on contrast-enhanced MRI). Final surgical pathology confirmed pCR of the primary tumor.

For prediction of axillary nodal pCR (Fig. 4B, Table 7), post-therapy [^68^Ga]Ga-FAPI-04 SULmean showed the highest diagnostic performance (AUC, 0.89; 95% CI, 0.7765-1.000; sensitivity, 91.67%; specificity, 81.25%; accuracy, 84.09%), whereas Δ SULmax% was the best-performing [^18^F]FDG-derived parameter (AUC, 0.669; 95% CI, 0.4875-0.851; sensitivity, 83.33%; specificity, 53.13%; accuracy, 61.36%). DeLong testing showed that [^68^Ga]Ga-FAPI-04 had significantly higher discriminative performance than [^18^F]FDG for nodal pCR (P < 0.05). A representative case is shown in Figure 6.

**FIGURE 6.**
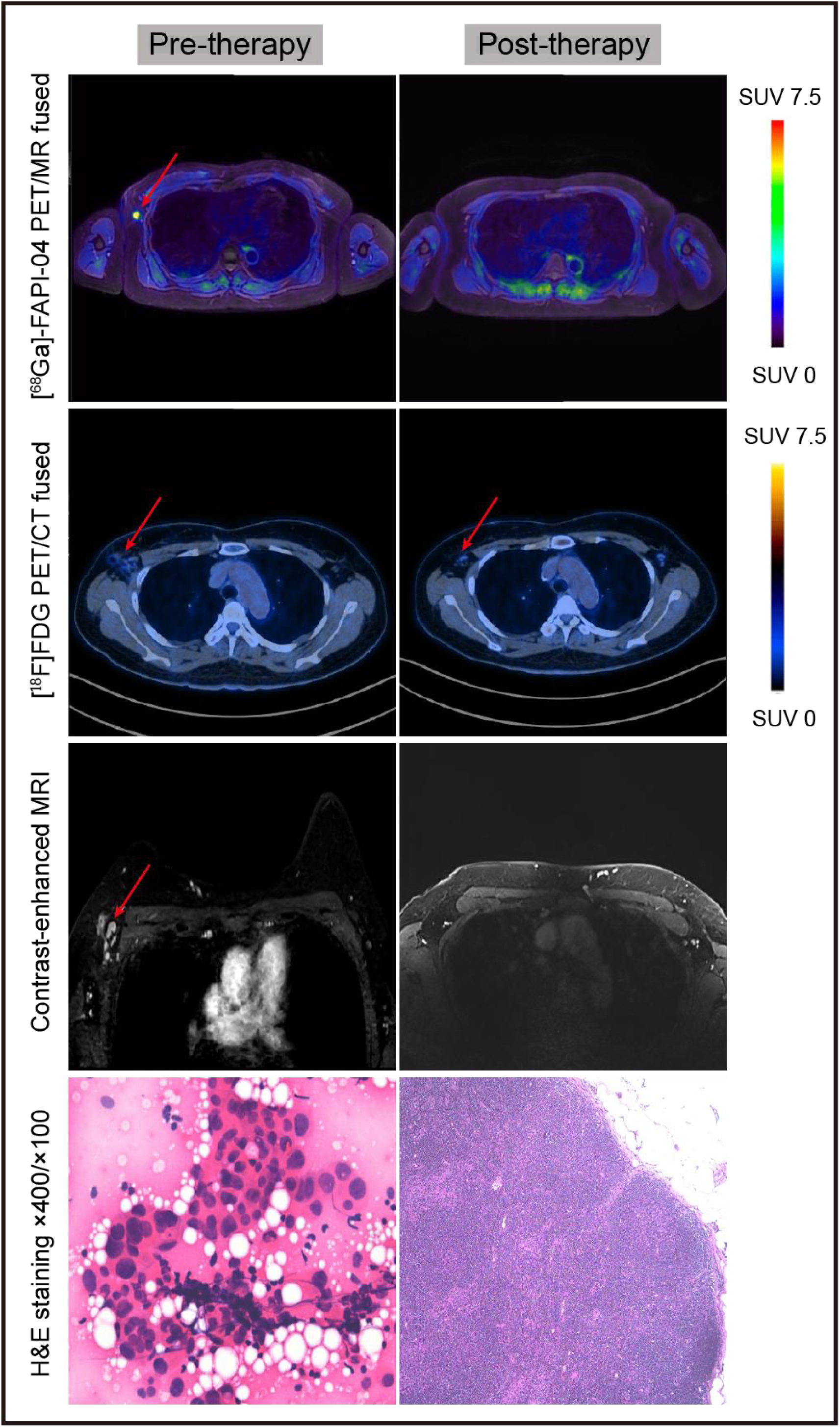
A woman with biopsy-proven axillary lymph node metastasis who achieved axillary pCR after NAC. Pre-therapy images obtained with three imaging modalities show metastatic axillary lymph nodes (red arrows), confirmed by cytologic biopsy. Post-therapy images show no abnormal tracer uptake in axillary lymph nodes on [^68^Ga]Ga-FAPI-04 PET, suggesting axillary pCR, whereas [^18^F]FDG PET shows residual uptake in a single lymph node (△SULmax%, 79.46%; below the cutoff value of 89.51%), indicating non-pCR. No abnormal enhancement is observed on contrast-enhanced MRI. Final surgical pathology confirms the absence of residual nodal metastasis.

**TABLE 7.** Diagnostic Performance of Imaging and Clinical Parameters for Prediction of pCR in Lymph Nodes.

| Parameter | Subparameter | Cutoff | AUC | P | 95%CI | Sensitivity(%) | Specificity(%) | Accuracy(%) |
| --- | --- | --- | --- | --- | --- | --- | --- | --- |
| Contrast-enhanced MRI in post-therapy |  | — | 0.77 | **0.01** | 0.563-0.977 | 75.00% | 78.95% | 77.78% |
| [<sup>68</sup>Ga]Ga-FAPI-04 | Post-therapy SULmean | 0.36 | 0.89 | **<0.01** | 0.7765-1 | 91.67% | 81.25% | 84.09% |
| [<sup>18</sup>F]FDG | Δ SULmax% | 89.51 | 0.669 | 0.08 | 0.4875-0.851 | 83.33% | 53.13% | 61.36% |
| NLR |  | 2.35 | 0.82 | **0.03** | 0.6231-1 | 100.00% | 61.54% | 73.68% |

AUC = Area Under the Curve.

### Association Between FAP Immunohistochemistry and ^68^Ga-FAPI Uptake

Fibroblast activation protein (FAP) immunohistochemical expression was assessed semiquantitatively using the predefined 4-tier stromal scoring system described in Methods. Paired pre-therapy and post-therapy specimens from 9 patients were analyzed for FAP staining. [^68^Ga]Ga-FAPI-04 SUVmax showed a strong positive correlation with stromal FAP expression (P < 0.001). FAP expression was markedly reduced in the original tumor bed of patients achieving pCR and decreased after chemotherapy in residual tumors, although levels remained higher than those observed in pCR tissues. Representative images are shown in Figure 7.

**FIGURE 7.**
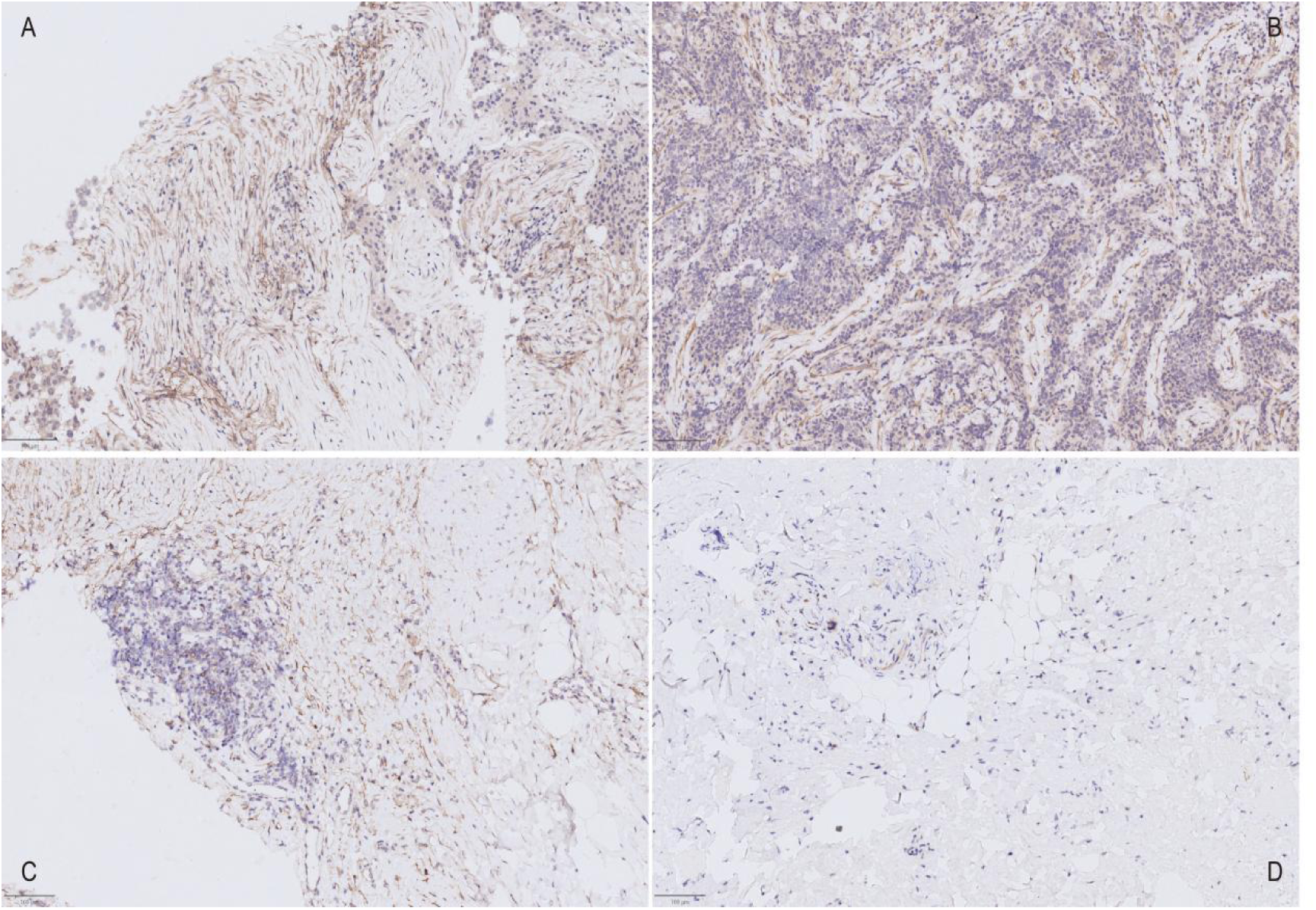
Comparison of FAP immunohistochemical staining in tumor tissues before and after chemotherapy in 2 patients. (A and C) Patient 1; (B and D) patient 2. (A and B) Pre-therapy images show high [^68^Ga]Ga-FAPI-04 uptake in patient 1 (SUVmax, 21.79; immunohistochemical score, 3) and moderate uptake in patient 2 (SUVmax, 8.8; score, 1). (C and D) Post-therapy images show reduced uptake in patient 1 without achieving pCR (SUVmax, 8.53; score, 1) and no visible tracer uptake in patient 2 with pCR (score, 0).

### Subgroup Analysis for Diagnostic Performance

Patients were stratified according to HER2 status to explore subgroup-specific diagnostic performance for primary-lesion pCR (Supplemental Table 7; Supplemental Fig. 3; Table 8). In HER2-positive patients, baseline [^68^Ga]Ga-FAPI-04 SUVmean was the optimal FAPI-derived parameter (AUC, 0.86), whereas post-therapy [^18^F]FDG MTV was the best-performing [^18^F]FDG parameter (AUC, 0.89); DeLong analysis showed no significant difference between the 2 tracers. In HER2-negative patients, no [^18^F]FDG parameter was significantly associated with pCR, whereas ΔSUVmax% on [^68^Ga]Ga-FAPI-04 showed excellent diagnostic performance (AUC, 0.94). Within subgroup-specific comparison families, the nominally significant HER2-positive and HER2-negative associations remained significant after false discovery rate correction; however, these subgroup findings should still be interpreted as exploratory because of the limited sample size (Supplemental Table 8).

**SUPPLEMENTAL TABLE 7.**
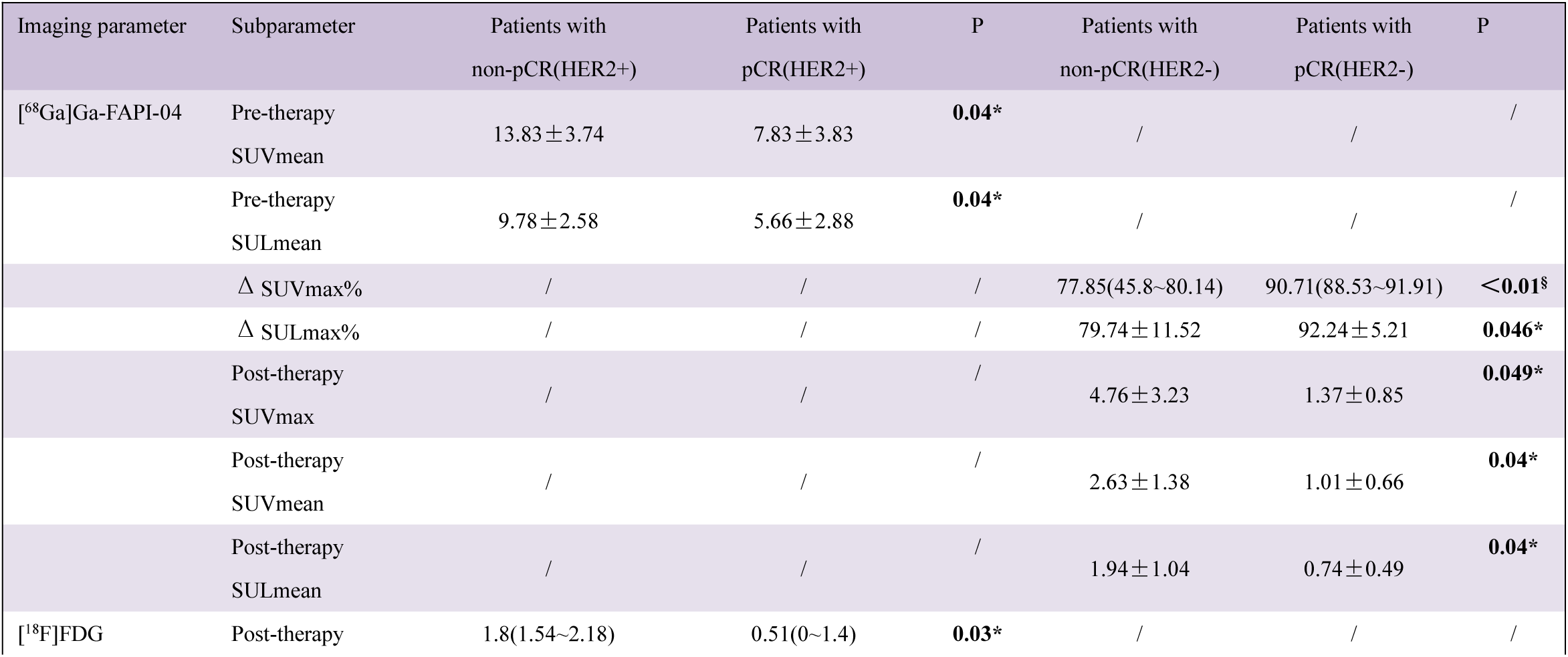
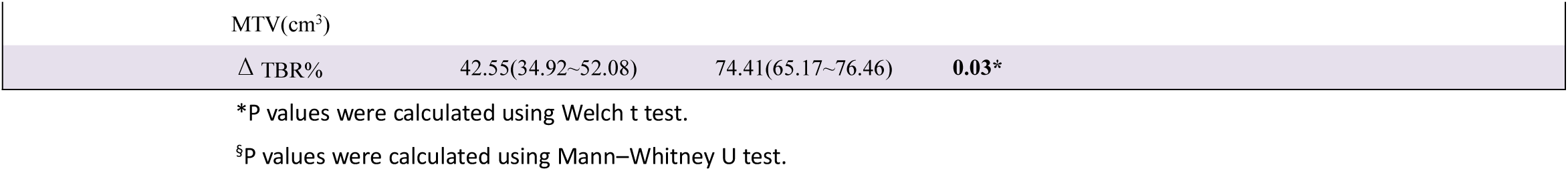
Subgroup Analysis of PET Characteristics According to Pathologic Response in Primary Lesions (Stratified by HER2 Expression)

**TABLE 8.**
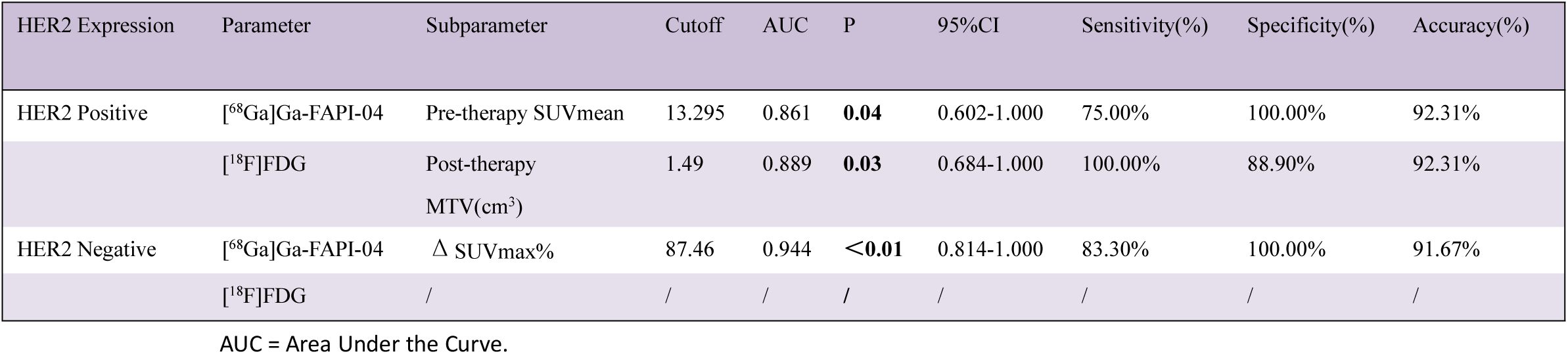
Subgroup Analysis of Diagnostic Performance of Imaging Parameters for Prediction of pCR in Primary Lesions (Stratified by HER2 Expression)

| HER2 Expression | Parameter | Subparameter | Cutoff | AUC | P | 95%CI | Sensitivity(%) | Specificity(%) | Accuracy(%) |
| --- | --- | --- | --- | --- | --- | --- | --- | --- | --- |
| HER2 Positive | [<sup>68</sup>Ga]Ga-FAPI-04 | Pre-therapy SUVmean | 13.295 | 0.861 | **0.04** | 0.602-1.000 | 75.00% | 100.00% | 92.31% |
|  | [<sup>18</sup>F]FDG | Post-therapy MTV(cm<sup>3</sup>) | 1.49 | 0.889 | **0.03** | 0.684-1.000 | 100.00% | 88.90% | 92.31% |
| HER2 Negative | [<sup>68</sup>Ga]Ga-FAPI-04 | $\Delta$ SUVmax% | 87.46 | 0.944 | **<0.01** | 0.814-1.000 | 83.30% | 100.00% | 91.67% |
|  | [<sup>18</sup>F]FDG | / | / | / | / | / | / | / | / |

AUC = Area Under the Curve.

**SUPPLEMENTAL FIGURE 3.**
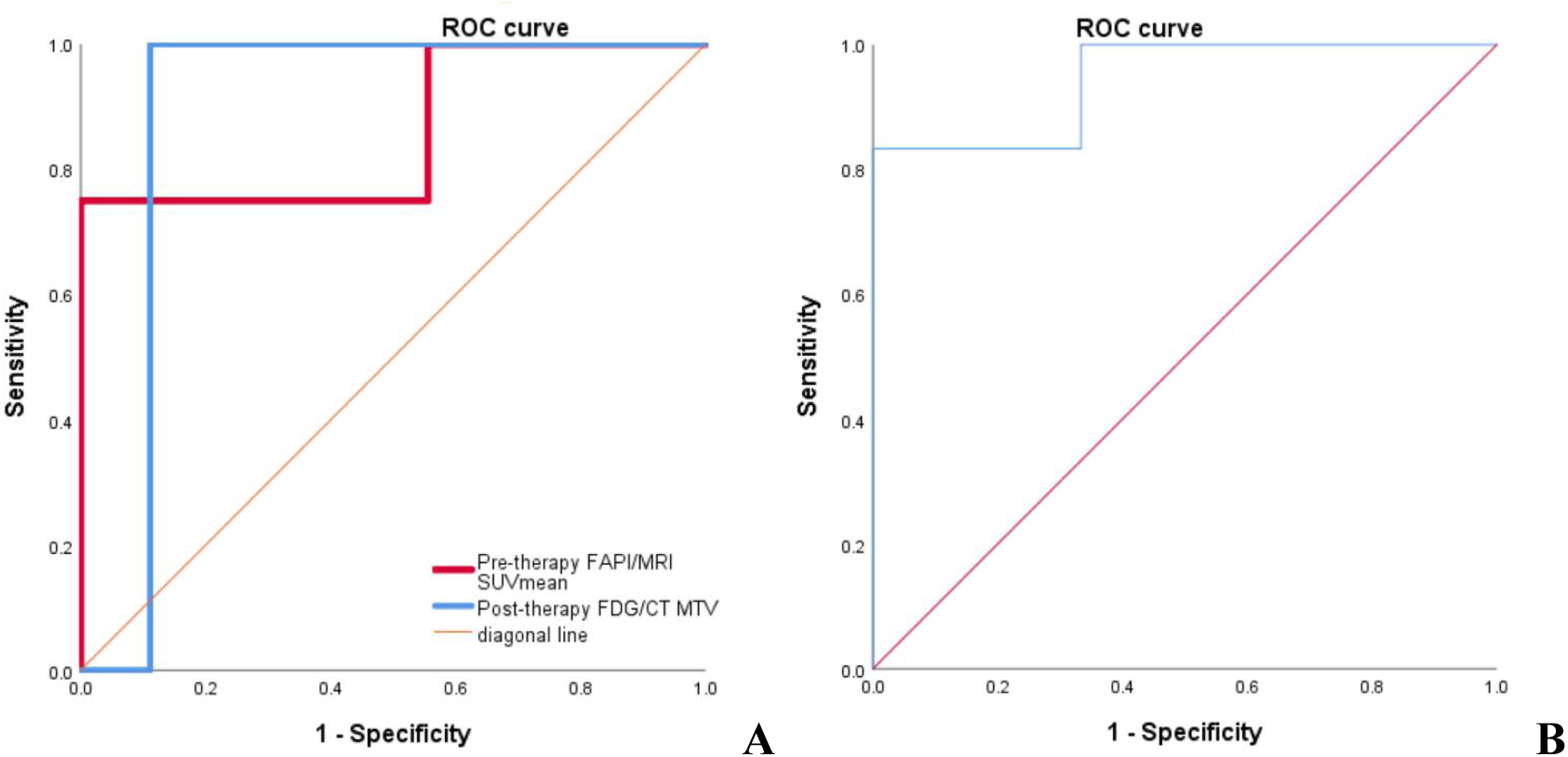
Receiver-operating-characteristic (ROC) curves of the optimal imaging parameters for prediction of pCR in HER2-defined subgroups. A: baseline [^68^Ga]Ga-FAPI-04 SUVmean and post-therapy [^18^F]FDG MTV in the HER2-positive subgroup. B: Δ SUVmax% on [^68^Ga]Ga-FAPI-04 PET in the HER2-negative subgroup.

**SUPPLEMENTAL TABLE 8.**
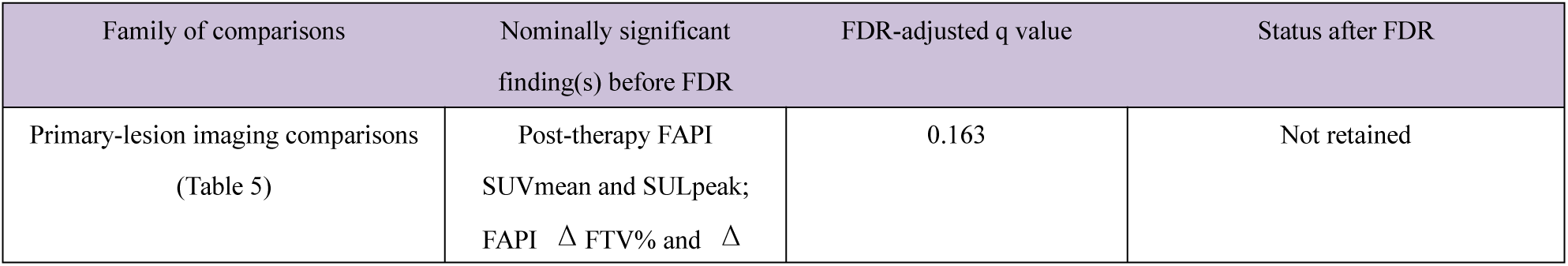
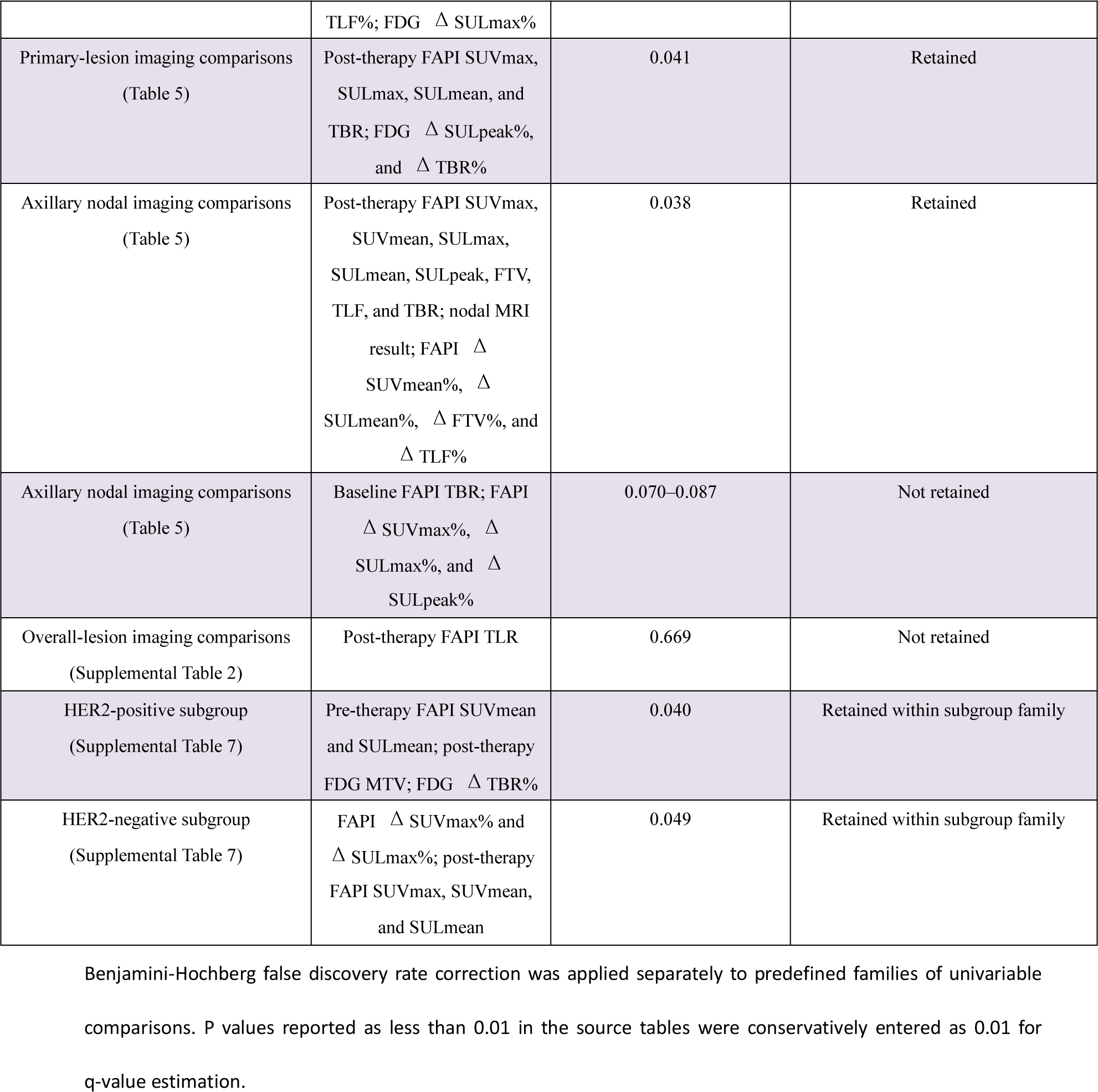
Summary of Benjamini-Hochberg False Discovery Rate Correction for Exploratory Univariable Analyses.

## DISCUSSION

Accurate identification of pathologic complete response (pCR) after neoadjuvant chemotherapy (NAC) has direct clinical relevance in breast cancer because it informs prognosis and may influence surgical decision-making, particularly in the axilla (2,3). In this prospective head-to-head study, post-therapy [^68^Ga]Ga-FAPI-04 PET showed the strongest association with pathologic response among the three imaging modalities evaluated. Specifically, post-therapy SUVmax was the optimal parameter for assessing response in primary lesions, whereas post-therapy SULmean yielded the highest diagnostic performance for axillary lymph nodes. In comparison, the best [^18^F]FDG-derived metrics showed only moderate discriminative ability, and contrast-enhanced MRI, although clinically useful, provided less diagnostic information than the strongest FAPI-derived parameters. These findings suggest that FAP-targeted imaging may offer a biologically meaningful approach for response assessment after NAC, particularly when viable tumor burden is low but treatment-related stromal remodeling remains active (8,9).

Our results are consistent with the growing body of evidence supporting FAPI-based imaging in breast cancer. Backhaus et al. reported promising initial results using ^68^Ga-FAPI-46 PET/MRI for response assessment after NAC, showing excellent correlation between follow-up FAPI uptake and pathologic response status (11). Systematic reviews focusing on breast and gynecologic malignancies have similarly highlighted the potential of FAPI imaging for improving lesion conspicuity, tumor-to-background contrast, and treatment response evaluation (10). More recent clinical studies further indicate that FAP-targeted imaging may outperform FDG in selected breast cancer contexts, underscoring the biological relevance of stromal imaging (12,19).

A key strength of the present study is the separate evaluation of primary-tumor and axillary nodal response. For primary lesions, the best FAPI-derived parameter numerically exceeded the best [^18^F]FDG-derived parameter, although the difference did not reach statistical significance. This pattern may reflect the biologic complexity of the post-NAC tumor bed, where residual fibrosis, isolated tumor cells, and therapy-induced inflammatory changes can confound image interpretation across modalities. In contrast, for axillary lymph nodes, FAPI imaging showed clearer qualitative and quantitative separation than [^18^F]FDG. This finding is clinically relevant because axillary status after NAC remains a critical determinant of prognosis and guides locoregional management, whereas nodal restaging with conventional imaging remains challenging after NAC (18,20).

From a clinical perspective, the potential implications of our findings extend to both the breast primary lesion and the axilla. For the primary tumor, more accurate estimation of residual disease after NAC may improve preoperative planning by informing the feasibility of breast-conserving surgery, the anticipated extent of resection, and the interpretation of treatment response when selecting postoperative systemic therapy. In recent years, several studies have suggested that, in carefully selected patients with excellent response to NAC, de-escalation of breast surgery—including increased use of breast-conserving approaches and, in exploratory settings, even omission of surgery—may be feasible, provided that residual disease can be reliably excluded (21). In this context, imaging modalities with improved accuracy in the post-therapy tumor bed are of particular interest. Although the advantage of [^68^Ga]Ga-FAPI-04 PET over [^18^F]FDG PET in the primary lesion was not statistically definitive in this cohort, its numerically higher diagnostic performance suggests that stromal-targeted imaging may provide clinically useful complementary information in the post-therapy tumor bed, where fibrosis, inflammation, and scattered residual disease can complicate conventional assessment. In parallel, the stronger performance observed for axillary nodal response is particularly relevant because residual nodal disease after NAC directly affects axillary surgical planning and subsequent locoregional and systemic treatment decisions. Notably, ongoing efforts toward axillary de-escalation, such as the selective omission of axillary lymph node dissection in patients with good nodal response, also rely heavily on accurate assessment of residual nodal disease (22,23). Taken together, these findings suggest that [^68^Ga]Ga-FAPI-04 PET may contribute to a more integrated preoperative assessment of residual disease burden in both the breast and axilla. However, these results should currently be regarded as complementary rather than practice-changing, and they are not sufficient to support omission of surgical pathologic evaluation.

The favorable performance of FAPI imaging in this context is biologically plausible and further supported by our immunohistochemical analyses. In the paired tissue subset, stromal FAP expression positively correlated with [^68^Ga]Ga-FAPI-04 SUVmax, and lesions achieving pCR exhibited minimal residual FAP expression in the original tumor bed. These observations align with current understanding that FAP-expressing cancer-associated fibroblasts play pivotal roles in extracellular matrix remodeling, immune modulation, and treatment resistance in breast cancer (9,24). If NAC effectively eliminates invasive tumor cells and suppresses stromal activation, both tissue FAP expression and FAPI tracer uptake would be expected to decline substantially—findings that were confirmed in our cohort.

MRI remained clinically valuable in our study, demonstrating significant predictive utility for both breast and nodal pCR, although its performance was inferior to that of the strongest FAPI-derived metrics. These findings are consistent with previous reports indicating that MRI is effective for monitoring response to NAC (4,25). However, MRI remains prone to false-positive interpretation because of fibrotic change, inflammatory reaction, and residual nonmass enhancement (5). The [^18^F]FDG PET results also warrant cautious interpretation. Although no post-therapy [^18^F]FDG parameter significantly predicted pCR in the primary lesion on univariable analysis, dynamic [^18^F]FDG markers such as ΔTBR% retained moderate diagnostic performance, consistent with prior evidence supporting serial [^18^F]FDG-based metrics for pCR prediction (6,26). [^18^F]FDG PET continues to play an important role in response-adapted strategies, particularly in HER2-positive disease (7,27). Exploratory data from the PHERGain trial also highlight the complementary value of [^18^F]FDG PET relative to breast MRI for predicting pCR and stratifying outcomes in early-stage HER2-positive disease (28). Nevertheless, compared with [^18^F]FDG, FAPI PET may provide higher contrast for detecting residual stromal activity and may be less influenced by biologic and physiologic confounders related to glucose metabolism.

The exploratory subgroup analysis according to HER2 status is noteworthy. In HER2-positive tumors, FAPI and [^18^F]FDG showed comparable discriminative ability for predicting primary-lesion pCR. In contrast, in HER2-negative disease, no [^18^F]FDG parameter was significantly associated with pCR, whereas ΔSUVmax% derived from FAPI PET showed excellent performance. Because of the limited sample size, these subgroup findings should be regarded as hypothesis-generating rather than practice-changing. Even so, they are consistent with prior observations that the predictive accuracy of [^18^F]FDG PET varies across molecular subtypes and treatment contexts (26). These data suggest that stromal-targeted imaging may be particularly useful in subgroups for which tumor-cell glucose metabolism is not a stable indicator of treatment response.

This study has several limitations. First, the sample size was modest and enrollment was conducted at a single center, which limits statistical power and generalizability. Second, the combined use of patient-level and lesion- or compartment-level analyses may introduce statistical dependence and may underestimate variability because observations within the same patient are not fully independent. Third, the primary-lesion analysis should be interpreted cautiously because the lesion-level sample size was small and the primary-lesion pCR rate was relatively high in this cohort (15/25, 60.0%), leaving only 10 non-pCR lesions for comparison; this class distribution may have reduced the precision of the primary-lesion estimates and partly limited our ability to demonstrate clearer statistical separation between imaging modalities. Fourth, subgroup analyses should be regarded as exploratory because of the small numbers and biologic heterogeneity. Fifth, MRI assessments were based on routine clinical interpretation rather than standardized fully quantitative multiparametric or radiomics approaches. Despite these limitations, the prospective design, direct multimodal comparison, dedicated nodal evaluation, and paired immunohistochemical correlation enhance the clinical relevance of the findings.

Overall, our data support post-therapy [^68^Ga]Ga-FAPI-04 PET as a promising imaging biomarker for assessing treatment response after NAC in breast cancer, particularly for evaluating axillary nodal clearance while also providing potentially useful complementary information for the primary tumor bed. Larger multicenter studies are needed to confirm the optimal imaging time point and quantitative thresholds and to determine whether integration of FAPI PET with MRI, molecular subtyping, and circulating biomarkers can improve personalized surgical and systemic treatment decision-making.

## CONCLUSION

In patients with breast cancer undergoing NAC, post-therapy [^68^Ga]Ga-FAPI-04 PET showed promising performance for predicting pathologic response. For primary breast tumors, [^68^Ga]Ga-FAPI-04 PET showed numerically higher diagnostic performance than [^18^F]FDG PET, whereas for axillary lymph nodes it showed significantly higher discriminative performance in this cohort. The concordance between FAPI uptake and stromal FAP expression supports the biologic validity of this imaging approach. Accordingly, [^68^Ga]Ga-FAPI-04 PET may serve as a valuable adjunct for post-NAC response assessment in the breast.

## Clinical Trial Registration

ClinicalTrials.gov, NCT07553741.

## Ethics Approval

This study was approved by the Ethics Committee of Union Hospital, Tongji Medical College, Huazhong University of Science and Technology (registration number: UHCT250235).

## Funding

None

## Competing Interests

The authors declare no competing interests.

## Data Availability

Deidentified data are available from the corresponding author upon reasonable request and subject to institutional and ethical approval.

## Author Contributions

Yuming Luo: Conceptualization, methodology, study implementation, investigation, data curation, formal analysis, visualization, project administration, writing—original draft, and writing—review and editing.

Xiao Zhang: Methodology, PET/MRI acquisition, image interpretation, imaging resources, investigation, visualization, and writing—review and editing.

Runze Li: Pathologic assessment, immunohistochemical analysis, validation, and writing—review and editing.

Yalan Zeng, Yu Zhao, Lei Li, Bei Qian, Yunxiao Xiao, Yaqi Zhao, Silin Xu, Qin Yang, Huanwei Zhang and Hengyu Chen: Patient recruitment, clinical data collection, investigation, data verification, and writing—review and editing.

Mengting Li: PET/MRI acquisition, image processing, image interpretation, investigation, and writing—review and editing.

Chong Lu: Conceptualization, resources, supervision, project administration, and writing—review and editing.

Xiaoli Lan: Conceptualization, funding acquisition, PET/MRI resources, imaging protocol supervision, resources, supervision, and writing—review and editing.

Chunping Liu: Conceptualization, supervision, project administration, ethics oversight, data governance, resources, and writing—review and editing.

All authors reviewed and approved the final version of the manuscript.

## DISCLOSURE

No potential conflict of interest relevant to this article was reported.

## Notes

### Competing Interest Statement

The authors have declared no competing interest.

### Clinical Trial

NCT07553741

