## Supplemental Tables for "Prospective Comparison of [^68^Ga]Ga-FAPI-04 PET, [^18^F]FDG PET, and Contrast-Enhanced MRI for Predicting Pathologic Response after Neoadjuvant Chemotherapy in Breast Cancer"

### Supplemental Table 1

#### Clinical Characteristics of Patients According to All Lesions Pathologic Response

| Characteristic | Patients with non-pCR | Patients with pCR | P |
|---|---|---|---|
| Age(y) | 52.2±8.6 | 47.21±9.96 | 0.20* |
| BMI | 24.34±2.44 | 23.69±1.92 | 0.50* |
| Menstrual state | | | 1† |
| Postmenopause | 4(44.44%) | 5(55.56%) | |
| Premenopause | 6(40.00%) | 9(60.00%) | |
| Clinical T category | | | 0.71# |
| cT0 | 1(100.00%) | 0(0.00%) | |
| cT1 | 1(50.00%) | 1(50.00%) | |
| cT2 | 7(41.18%) | 10(58.82%) | |
| cT3 | 1(25.00%) | 3(75.00%) | |
| Clinical N category | | | 0.70# |
| cN0 | 2(40.00%) | 3(60.00%) | |
| cN1 | 6(46.15%) | 7(53.85%) | |
| cN2 | 0(0.00%) | 2(100.00%) | |
| cN3 | 2(50.00%) | 2(50.00%) | |
| AJCC Clinical stage | | | 1† |
| Stage II | 7(41.18%) | 10(58.82%) | |
| Stage III | 3(42.86%) | 4(57.14%) | |
| Ki-67 | | | 0.24† |
| High expression | 3(27.27%) | 8(72.73%) | |
| Low expression | 7(53.85%) | 6(46.15%) | |
| Histological grading | | | 1† |
| G2 | 3(42.86%) | 4(57.14%) | |
| G3 | 7(41.18%) | 10(58.82%) | |
| Molecular subtype | | | 0.46# |
| Luminal | 3(75.00%) | 1(25.00%) | |
| HER2 positive | 4(33.33%) | 8(66.67%) | |
| Triple-negative | 3(37.50%) | 5(62.50%) | |
| Postoperative | | | |
| NLR | 2.41(2.26~3.35) | 2.11(1.79~2.94) | 0.25§ |
| PLR | 154.2(106.29~206.05) | 126.4(104.83~177.66) | 0.58§ |
| LMR | 2.85(1.83~4.06) | 3.14(2.29~3.59) | 0.71§ |
| SII | 493.84(302.26~993.57) | 399.14(263.63~626.17) | 0.71§ |
| HALP score | 30.82(22.6~40.18) | 36.64(30.87~46.53) | 0.38§ |
| PNI | 424.39±47.32 | 445.56±28.32 | 0.23* |

*P values were calculated using Welch t test.

#P values were calculated using likelihood ratio test.

†P values were calculated using Fisher–Freeman–Halton exact test.

§P values were calculated using Mann–Whitney U test.

NLR = Neutrophil - Lymphocyte Ratio;PLR= Platelet - Lymphocyte Ratio;LMR = Lymphocyte - Monocyte Ratio;SII = Systemic

Immune Inflammation Index;PNI = Prognostic Nutritional Index.

Qualitative data are number and percentage; continuous data are mean ± SD or median and 25th percentiles ~ 75th percentiles.

### Supplemental Table 2

#### PET Characteristics of Patients According to Pathologic Response

| Imaging parameter | Subparameter | Patients with non-pCR | Patients with pCR | P |
|---|---|---|---|---|
| Pretherapy | | | | |
| [68Ga]Ga-FAPI-04 | SUVmax (g/mL) | 16.66±6.44 | 16.85±6.23 | 0.94* |
| | SUVpeak (g/mL) | 12.88±5.65 | 13.4±5.2 | 0.82* |
| | SULmax (g/mL) | 9.77(8.99~15.4) | 12.5(9.64~16.24) | 0.82§ |
| | SULpeak (g/mL) | 8.96±4.11 | 9.52±3.95 | 0.73* |
| | FTV(cm3) | 19.57(12.68~32.74) | 23.13(10.15~33.23) | 0.91§ |
| | TLF(g/mL3cm3) | 122.83(72.94~342.2) | 161.46(74.53~295.62) | 0.91§ |
| | TLR | 16.39±6.35 | 18.29±9.66 | 0.57* |
| [18F]FDG | SUVmax (g/mL) | 10.58±4.27 | 13.39±7.34 | 0.26* |
| | SUVpeak (g/mL) | 5.6(5~9.27) | 9.83(6.54~13.34) | 0.42§ |
| | SULmax (g/mL) | 7.6±3.03 | 9.63±5.21 | 0.25* |
| | SULpeak (g/mL) | 4.27(3.49~6.4) | 6.39(4.87~9.52) | 0.38§ |
| | MTV(cm3) | 16.24(8.89~31.95) | 13.7(9.68~25.41) | 0.86§ |
| | TLG(g/mL3cm3) | 55.15(31.46~196.05) | 110.32(39.69~152.22) | 0.73§ |
| | TLR | 4.06±1.94 | 4.84±2.8 | 0.43* |
| Posttherapy | | | | |
| [68Ga]Ga-FAPI-04 | SUVmax (g/mL) | 2.61(2.26~6.37) | 2.11(1.04~2.52) | 0.08§ |
| | SUVpeak (g/mL) | 1.68(1.42~3.52) | 1.37(0.7~1.62) | 0.09§ |
| | SULmax (g/mL) | 1.86(1.69~4.7) | 1.37(0.73~1.85) | 0.08§ |
| | SULpeak (g/mL) | 1.2(1.01~2.52) | 1.02(0.49~1.22) | 0.12§ |
| | FTV(cm3) | 1.65(1.11~3.12) | 1.72(0~2.19) | 0.56§ |
| | TLF(g/mL3cm3) | 2.63(2.06~7.58) | 2.58(0~3.16) | 0.28§ |
| | TLR | 2.97±2.3 | 1.36±0.69 | **0.04*** |
| [18F]FDG | SUVmax (g/mL) | 1.69(1.62~2.26) | 1.65(1.43~2.09) | 0.43§ |
| | SUVpeak (g/mL) | 1.23(1.17~1.46) | 1.19(0.87~1.37) | 0.22§ |
| | SULmax (g/mL) | 1.21(1.18~1.63) | 1.17(1.04~1.51) | 0.37§ |
| | SULpeak (g/mL) | 0.91(0.85~1.04) | 0.88(0.6~0.97) | 0.22§ |
| | MTV(cm3) | 2.36(1.71~3.15) | 2.3(1.16~3.52) | 0.56§ |
| | TLG(g/mL3cm3) | 2.96(2.62~4.5) | 2.96(1.18~5) | 0.54§ |
| | TLR | 0.59(0.57~0.68) | 0.57(0.52~0.69) | 0.37§ |
| Pre- to posttherapy percentage change | | | | |
| [68Ga]Ga-FAPI-04 | △SUVmax% | 82.47(56.46~89.49) | 86.86(82.33~90.27) | 0.30§ |

| | | | | |
|---|---|---|---|---|
| | △SUVpeak% | 86.68(70.03~91.52) | 89.25(83.78~94.74) | 0.33§ |
| | △SULmax% | 81.94(54.03~89.39) | 86.71(82.44~90.78) | 0.25§ |
| | △SULpeak% | 86.96(68.4~91.42) | 87.44(84.26~94.98) | 0.39§ |
| | △FTV% | 87.36±11.97 | 92.65±7.14 | 0.22* |
| | △TLF% | 98.5(92.59~99.28) | 98.61(96.57~100) | 0.23§ |
| | △TLR% | 91.11(67.56~92.57) | 90.87(89.02~94.98) | 0.46§ |
| [18F]FDG | △SUVmax% | 76.27(72.66~87.11) | 85.76(77.87~89.09) | 0.33§ |
| | △SUVpeak% | 77.15(71.79~86.38) | 87.95(81.89~92.15) | 0.17§ |
| | △SULmax% | 76.69(72.41~86.92) | 86.12(77.86~89.14) | 0.33§ |
| | △SULpeak% | 76.12±13.42 | 84.55±10.95 | 0.11* |
| | △MTV% | 84.8(76.07~92.36) | 90.32(74.3~96.84) | 0.86§ |
| | △TLG% | 96.63(89.65~98.06) | 96.1(95.07~99.38) | 0.32§ |
| | △TLR% | 78.42(73.75~87.96) | 85.96(79.08~88.58) | 0.57§ |

*P values were calculated using Welch t test.

§P values were calculated using Mann–Whitney U test.

Continuous data are mean ± SD or median and 25th percentiles ~ 75th percentiles.

#### Supplemental Table 3

Diagnostic Performance of Imaging Parameters for All Lesions pCR

| Parameter | Subparameter | Cutoff | AUC | P | 95%CI | Sensitivity(%) | Specificity(%) | Accuracy(%) |
|---|---|---|---|---|---|---|---|---|
| [68Ga]Ga-FAPI-04 | Posttherapy TLR | 2.397 | 0.699 | **0.04** | 0.4782-0.9204 | 45.45% | 84.62% | 66.67% |

AUC = Area Under Curve.

#### Supplemental Table 4

PET Characteristics of Patients According to Primary Lesions HER2 Expression

| Imaging parameter | Subparameter | Patients with HER2(-) | Patients with HER2(+) | P |
|---|---|---|---|---|
| [68Ga]Ga-FAPI-04 | Pretherapy SUVmean(g/mL) | 6.5±2.18 | 9.67±4.65 | **0.041*** |
| | Pretherapy SULmean(g/mL) | 4.7±1.61 | 6.93±3.33 | **0.046*** |
| [18F]FDG | / | / | / | / |

*P values were calculated using Welch t test.

#### Supplemental Table 7

Subgroup Analysis for PET Characteristics of Patients According to Primary Lesions Pathologic Response (Based on HER2 Expression)

| Imaging parameter | Subparameter | Patients with non-pCR(HER2+) | Patients with pCR(HER2+) | P | Patients with non-pCR(HER2-) | Patients with pCR(HER2-) | P |
|---|---|---|---|---|---|---|---|
| [$^{68}$Ga]Ga-FAPI-04 | Pretherapy SUVmean(g/mL) | 13.83±3.74 | 7.83±3.83 | **0.04*** | / | / | / |
| | Pretherapy SULmean(g/mL) | 9.78±2.58 | 5.66±2.88 | **0.04*** | / | / | / |
| | △SUVmax% | / | / | / | 77.85(45.8~80.14) | 90.71(88.53~91.91) | **＜0.01§** |
| | △SULmax% | / | / | / | 79.74±11.52 | 92.24±5.21 | **0.046*** |
| | Posttherapy SUVmax (g/mL) | / | / | / | 4.76±3.23 | 1.37±0.85 | **0.049*** |
| | Posttherapy SUVmean (g/mL) | / | / | / | 2.63±1.38 | 1.01±0.66 | **0.04*** |
| | Posttherapy SULmean (g/mL) | / | / | / | 1.94±1.04 | 0.74±0.49 | **0.04*** |
| [$^{18}$F]FDG | Posttherapy MTV(cm3) | 1.8(1.54~2.18) | 0.51(0~1.4) | **0.03*** | / | / | / |
| | △TBR% | 42.55(34.92~52.08) | 74.41(65.17~76.46) | **0.03*** | | | |

*P values were calculated using Welch t test.

§P values were calculated using Mann–Whitney U test.

#### SUPPLEMENTAL TABLE 8

Summary of Benjamini-Hochberg False Discovery Rate Correction for Exploratory Univariable Analyses

| Family of comparisons | Nominally significant finding(s) before FDR | FDR-adjusted q value | Status after FDR |
|---|---|---|---|
| Primary-lesion imaging comparisons (Table 5) | post-therapy FAPI SUVmean and SULpeak; FAPI Δ FTV% and Δ TLF%; FDG Δ SULmax% | 0.163 | Not retained |
| Primary-lesion imaging comparisons (Table 5) | post-therapy FAPI SUVmax, SULmax, SULmean, and TBR; FDG Δ SULpeak%, and Δ TBR% | 0.041 | Retained |
| Axillary nodal imaging comparisons (Table 5) | post-therapy FAPI SUVmax, SUVmean, SULmax, SULmean, SULpeak, FTV, TLF, and TBR; nodal MRI result; FAPI Δ SUVmean%, Δ SULmean%, Δ FTV%, and | 0.038 | Retained |

| | | | |
|---|---|---|---|
| | Δ TLF% | | |
| Axillary nodal imaging comparisons (Table 5) | Baseline FAPI TBR; FAPI Δ SUVmax%, Δ SULmax%, and Δ SULpeak% | 0.070–0.087 | Not retained |
| Overall-lesion imaging comparisons (Supplemental Table 2) | post-therapy FAPI TLR | 0.669 | Not retained |
| HER2-positive subgroup (Supplemental Table 7) | pre-therapy FAPI SUVmean and SULmean; post-therapy FDG MTV; FDG Δ TBR% | 0.040 | Retained within subgroup family |
| HER2-negative subgroup (Supplemental Table 7) | FAPI Δ SUVmax% and Δ SULmax%; post-therapy FAPI SUVmax, SUVmean, and SULmean | 0.049 | Retained within subgroup family |

Benjamini-Hochberg false discovery rate correction was applied separately to predefined families of univariable comparisons. P values reported as less than 0.01 in the source tables were conservatively entered as 0.01 for q-value estimation.
