## Supplemental Figures for "Prospective Comparison of [^68^Ga]Ga-FAPI-04 PET, [^18^F]FDG PET, and Contrast-Enhanced MRI for Predicting Pathologic Response after Neoadjuvant Chemotherapy in Breast Cancer"

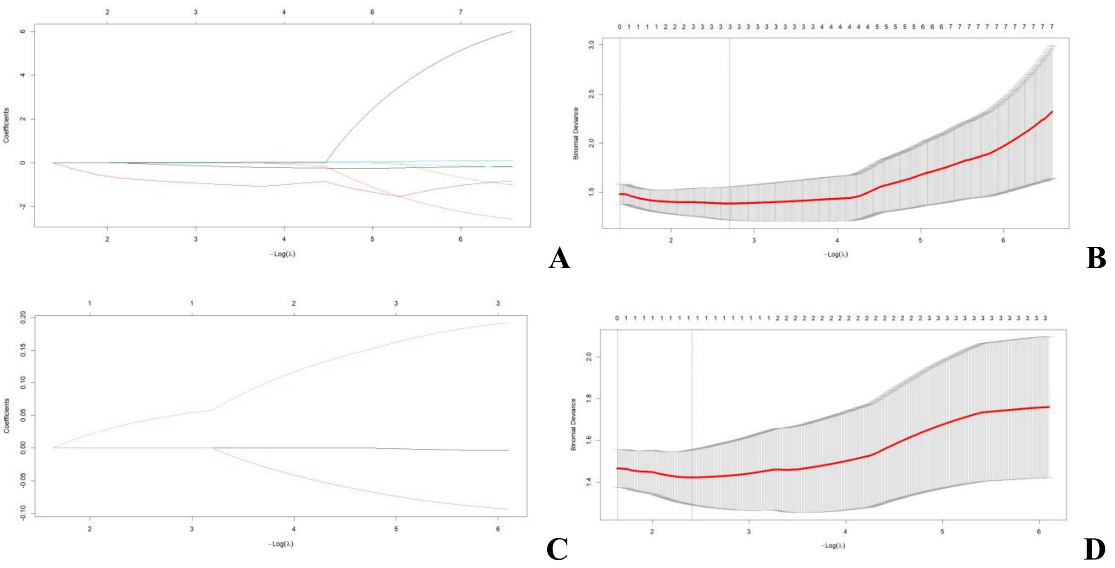


**SUPPLEMENTAL FIGURE 1.** LASSO coefficient profiles for primary-lesion analyses. A and B show the coefficient profile plot and cross-validation curve for [68Ga]Ga-FAPI-04, respectively. C and D show the coefficient profile plot and cross-validation curve for [18F]FDG, respectively. The optimal lambda was selected on the basis of minimum binomial deviance.

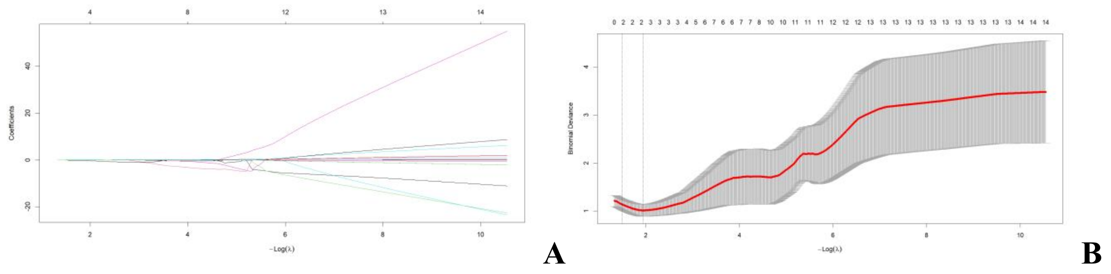


**SUPPLEMENTAL FIGURE 2.** LASSO coefficient profiles for axillary nodal analyses. A shows the coefficient profile plot. B shows the cross-validation curve. The optimal lambda was selected on the basis of minimum binomial deviance.

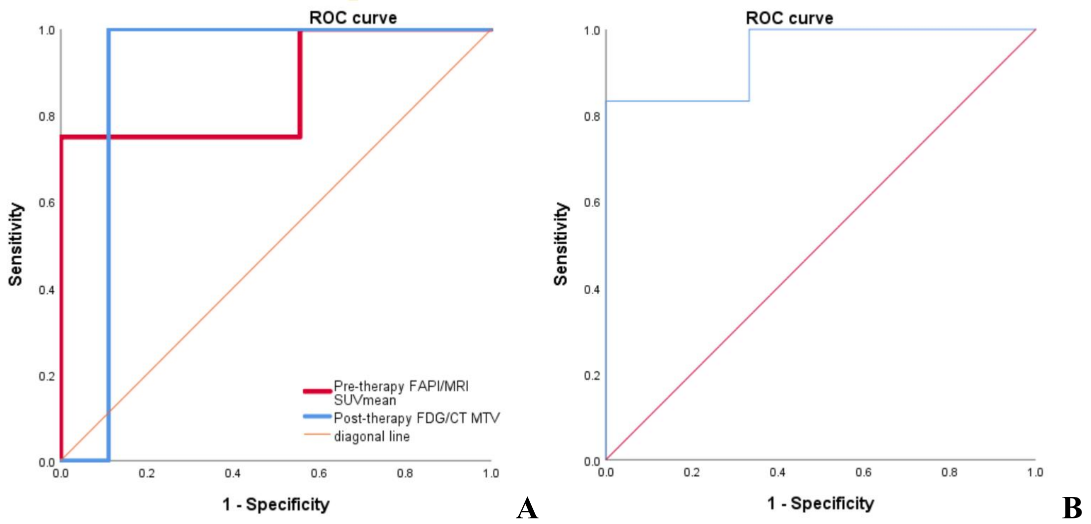


**SUPPLEMENTAL FIGURE 3.** Receiver-operating-characteristic (ROC) curves of the optimal imaging parameters for prediction of pCR in HER2-defined subgroups. A: baseline [$^{68}$Ga]Ga-FAPI-04 SUVmean and post-therapy [$^{18}$F]FDG MTV in the HER2-positive subgroup. B: △SUVmax% on [$^{68}$Ga]Ga-FAPI-04 PET in the HER2-negative subgroup.
